# Type 2 diabetes polygenic risk score demonstrates context-dependent effects and associations with type 2 diabetes-related risk factors and complications across diverse populations

**DOI:** 10.1101/2025.02.15.25322341

**Authors:** Boya Guo, Yanwei Cai, Daeeun Kim, Roelof A.J. Smit, Zhe Wang, Kruthika R. Iyer, Austin T Hilliard, Jeffrey Haessler, Ran Tao, K Alaine Broadaway, Yujie Wang, Nikita Pozdeyev, Frederik F. Stæger, Chaojie Yang, Brett Vanderwerff, Amit D. Patki, Lauren Stalbow, Meng Lin, Nicholas Rafaels, Jonathan Shortt, Laura Wiley, Maggie Stanislawski, Jack Pattee, Lea Davis, Peter S. Straub, Megan M. Shuey, Nancy J. Cox, Nanette R. Lee, Marit E. Jørgensen, Peter Bjerregaard, Christina Larsen, Torben Hansen, Ida Moltke, James B. Meigs, Daniel O. Stram, Xianyong Yin, Xiang Zhou, Kyong-Mi Chang, Shoa L. Clarke, Rodrigo Guarischi-Sousa, Joanna Lankester, Philip S Tsao, Steven Buyske, Mariaelisa Graff, Laura M. Raffield, Quan Sun, Lynne R. Wilkens, Christopher S. Carlson, Charles B. Easton, Simin Liu, JoAnn E. Manson, Loïc L. Marchand, Christopher A. Haiman, Karen L. Mohlke, Penny Gordon-Larsen, Anders Albrechtsen, Michael Boehnke, Stephen S. Rich, Ani Manichaikul, Jerome I. Rotter, Noha A. Yousri, Ryan M. Irvin, The biobank at the Colorado Center for Personalized Medicine, VA Million Veteran Program, The Population Architecture using Genomics and Epidemiology (PAGE) study, Chris Gignoux, Kari E. North, Ruth J.F. Loos, Themistocles L. Assimes, Ulrike Peters, Charles Kooperberg, Sridharan Raghavan, Heather M. Highland, Burcu F. Darst

**Affiliations:** Public Health Sciences, Fred Hutchinson Cancer Center, Seattle, WA, USA; Department of Epidemiology, Gillings School of Global Public Health, University of North Carolina at Chapel Hill, Chapel Hill, NC, USA; Department of Genetics, University of North Carolina at Chapel Hill, Chapel Hill, NC, USA; The Charles Bronfman Institute for Personalized Medicine, Icahn School of Medicine at Mount Sinai, New York, NY, USA; Data Science and Biotechnology, Gladstone Institutes, San Francisco, CA, USA; Department of Medicine, Division of Cardiovascular Medicine, Stanford University School of Medicine, Stanford, CA, USA; VA Palo Alto Health Care System, Palo Alto, CA, USA; Department of Biostatistics, Vanderbilt University Medical Center, Nashville, TN, USA; Vanderbilt Genetic Institute, Vanderbilt University of Medical Center, Nashville, TN, USA; Colorado Center for Personalized Medicine, University of Colorado Anschutz Medical Campus, Aurora, CO, USA; Department of Biology, Section for Computational and RNA Biology, University of Copenhagen, Copenhagen, Denmark; Department of Genome Sciences, University of Virginia, Charlottesville, VA, USA; Department of Biostatistics and Center for Statistical Genetics, University of Michigan, Ann Arbor, MI, USA; Department of Epidemiology, School of Public Health, University of Alabama at Birmingham, Birmingham, AL, USA; Department of Biostatistics and Informatics, Colorado School of Public Health, University of Colorado Anschutz Medical Campus, Aurora, CO, USA; Department of Medicine, Division of Data-Driven and Digital Medicine, Icahn School of Medicine at Mount Sinai, New York, NY, USA; Department of Medicine, Division of Genetic Medicine, Vanderbilt University of Medical Center, Nashville, TN, USA; USC-Office of Population Studies Foundation, Inc., University of San Carlos, Cebu City, Philippines; Steno Diabetes Center Greenland, Nuuk, Greenland; Centre for Public Health in Greenland, National Institute of Public Health, University of Southern Denmark, Denmark; Novo Nordisk Foundation Center for Basic Metabolic Research, Faculty of Health and Medical Sciences, University of Copenhagen, Copenhagen, Denmark; Department of Medicine, Mass General Brigham Healthcare System, Harvard Medical School, Boston, MA, USA; Department of Preventive Medicine, Keck School of Medicine, University of Southern California, Los Angeles, CA, USA; Department of Epidemiology, School of Public Health, Nanjing Medical University, Nanjing, China; Corporal Michael J. Crescenz VA Medical Center, Philadelphia, PA, USA; Department of Medicine, Division of Gastroenterology and Hepatology, University of Pennsylvania Perelman School of Medicine, Philadelphia, PA, USA; Department of Medicine, Stanford Prevention Research Center, Stanford University School of Medicine, Stanford, CA, USA; Department of Genetics, Rutgers University, Piscataway, NJ, USA; Department of Biostatistics, Gillings School of Global Public Health, University of North Carolina at Chapel Hill, Chapel Hill, NC, USA; Epidemiology Program, University of Hawaii Cancer Center, Honolulu, HI, USA; Family Medicine and Epidemiology, Warren Alpert Medical School and School of Public Health, Brown University, Pawtucket, RI, USA; Department of Epidemiology, School of Public Health, Brown University, Providence, RI, USA; Department of Nutrition, Gillings School of Global Public Health, University of North Carolina at Chapel Hill, Chapel Hill, NC, USA; Department of Pediatrics, The Institute for Translational Genomics and Population Sciences, The Lundquist Institute for Biomedical Innovation at Harbor-UCLA Medical Center, Torrance, CA, USA; College of Health and Life Sciences, Hamad Bin Khalifa University, Doha, Qatar; Computer and Systems Engineering, Alexandria University, Alexandria, Egypt; A list of consortium members can be found in the Supplementary Information; Department of Epidemiology, School of Public Health, University of Washington, Seattle, WA, USA; Department of Medicine, Division of General Internal Medicine, University of Colorado School of Medicine, Denver, CO, USA

## Abstract

Polygenic risk scores (PRS) hold prognostic value for identifying individuals at higher risk of type 2 diabetes (T2D). However, further characterization is needed to understand the generalizability of T2D PRS in diverse populations across various contexts. We characterized a multi-ancestry T2D PRS among 244,637 cases and 637,891 controls across eight populations from the Population Architecture Genomics and Epidemiology (PAGE) Study and 13 additional biobanks and cohorts. PRS performance was context dependent, with better performance in those who were younger, male, with a family history of T2D, without hypertension, and not obese or overweight. Additionally, the PRS was associated with various diabetes-related cardiometabolic traits and T2D complications, suggesting its utility for stratifying risk of complications and identifying shared genetic architecture between T2D and other diseases. These findings highlight the need to account for context when evaluating PRS as a tool for T2D risk prognostication and potentially generalizable associations of T2D PRS with diabetes-related traits despite differential performance in T2D prediction across diverse populations.

## Main

The rise in type 2 diabetes (T2D) prevalence is a major public health challenge which shows no signs of abating. Without sufficient efforts to combat this pandemic, it has been estimated that 11.3% of the global population will have diabetes by 2030^1^, at which time its economic burden is projected to be 2.2% of the global gross domestic product.^2^ These predictions provide an urgent incentive to improve diabetes risk prediction, stratification, and prevention.

An emerging and promising approach to quantify T2D risk is the use of polygenic risk scores (PRS), which model the aggregate effect of genetic variants found across the genome on a disease or trait of interest. Such scores, combined with non-genetic predictors, potentially hold utility for identifying individuals at high risk prior to disease onset and could guide positive health behavior changes, screening practices, or therapeutic interventions.^3–5^ These PRS have been facilitated by genome-wide association studies (GWAS) that have identified genetic variants conferring T2D susceptibility, with several recent T2D GWAS and fine mapping studies conducted across diverse populations enabling the development of multi-ancestry T2D PRS.^6–8^

Given the large number of risk factors for T2D, there is a clear need to further characterize these scores. Although previous studies have reported that interactions between genetic variants and certain lifestyle factors influence T2D risk in European ancestry populations^8–11^, little is known about whether the predictive accuracy and utility of T2D PRS is context-dependent and modified by individual characteristics and metabolic parameters across diverse populations. Moreover, it remains unclear whether PRS for T2D risk could hold prognostic value for diabetes-related complications across numerous organ systems in those with clinically overt disease. A recent investigation found that a T2D PRS was associated with T2D-related retinopathy, with modest evidence of association with chronic kidney disease, peripheral artery disease, and neuropathy in individuals who were genetically similar to European ancestry.^7^ A thorough investigation of these and other complications across diverse populations could provide biological insights into the pleiotropic nature and prognostic ability of genetic variants associated with T2D risk.

The Population Architecture using Genomics and Epidemiology (PAGE) Study, which consists of multiple large and diverse population-based studies, is uniquely situated to address these questions. In PAGE, we previously found that a multi-ancestry T2D PRS outperformed population-specific T2D PRS and explained up to 15.3% of familial relative risk of T2D across diverse populations.^8^ Here, after identifying a multi-ancestry T2D PRS with optimal performance in PAGE, we investigated whether the effect of a T2D PRS was context-dependent across PAGE and several independent biobanks and cohorts. To further understand the clinical implications of genetic risk of T2D on other diseases and conditions, we examined whether a T2D PRS was associated with T2D-related phenotypic risk factors and complications and additionally conducted phenome-wide association studies (PheWASs) across diverse populations. The goal of this investigation was to improve precision medicine in diabetes by better characterizing genetic risk of T2D and identifying individuals at greater risk of developing diabetes-related complications.

## Results

### Participants

An overview of the study design is provided in **Figure 1**. In total, 82,944 participants from PAGE and 835,241 participants from 13 additional biobanks and cohorts were included in this study: 244,637 prevalent T2D cases, 637,891 controls, and 35,657 individuals with prediabetes. This included individuals who self-reported as African, African American, or Black (AFR); American Indian, Alaskan Native, or Native American (AI); Asian American, East of Southeast Asian, or South Asian/Indian (ASN); White or European American (EUR); Greenlandic; Hispanic/Latino American (HIS); Middle Eastern, North African, or Qatari (MENAQ); and Native Hawaiian or Other Pacific Islander (NH) (**Table 1**, **Table S1, Methods, and Supplemental Information** for population descriptor details).

**Figure 1.**
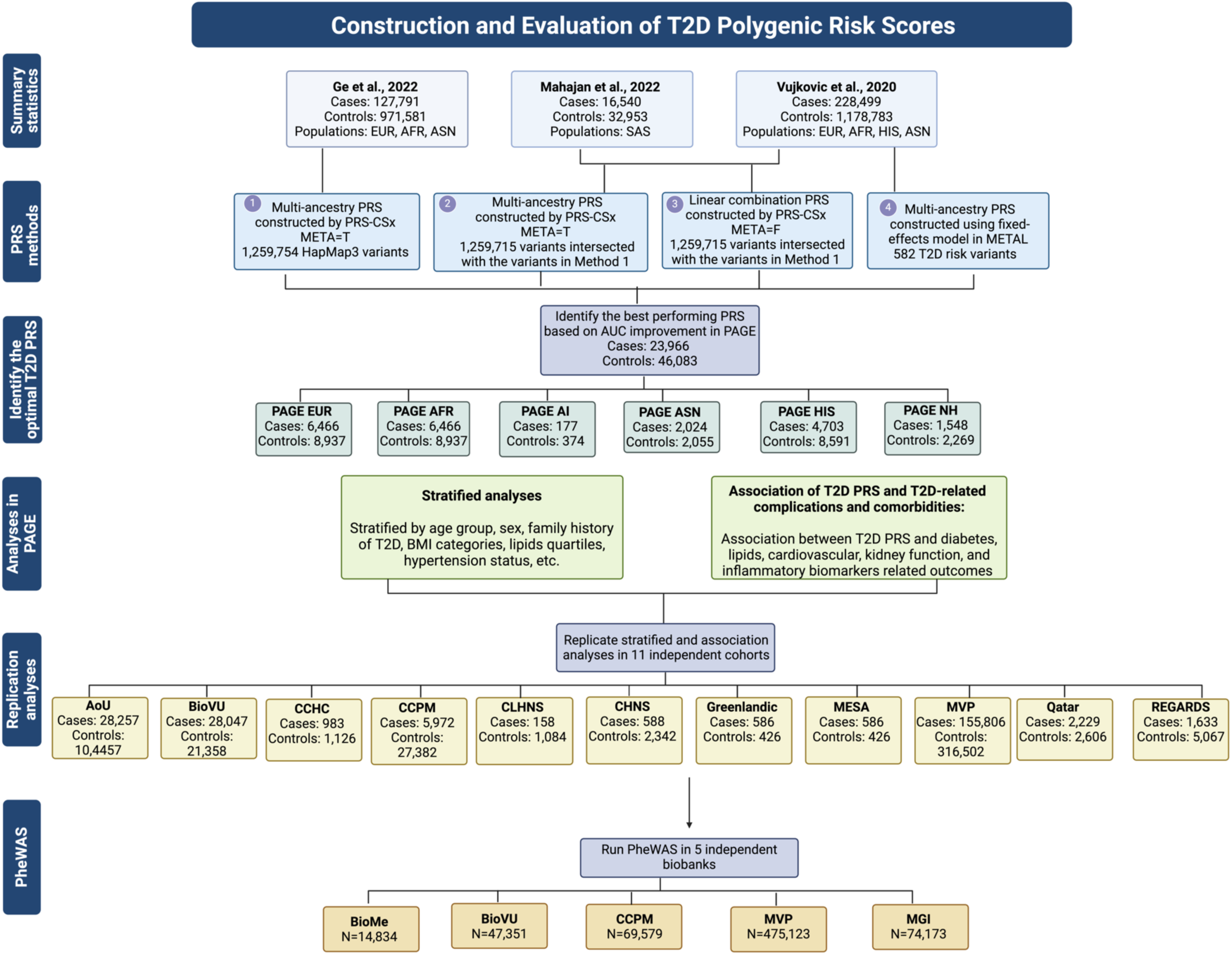
Workflow of T2D PRS construction and evaluation.

**Table 1.** Sample sizes of populations included by T2D status.

| **Population** | **T2D cases** | **T2D controls** | **Prediabetes** | **Total** |
| --- | --- | --- | --- | --- |
| AFR | 52,159 | 106,306 | 2,333 | 160,798 |
| AI | 177 | 374 | 91 | 642 |
| ASN | 6,999 | 17,660 | 3,134 | 27,793 |
| EUR | 148,230 | 441,879 | 18,126 | 608,235 |
| Greenlandic | 586 | 426 | 2,824 | 3,836 |
| HIS | 32,440 | 65,613 | 8,955 | 107,008 |
| MENAQ | 2,352 | 3,078 | 53 | 5,483 |
| NH | 1,694 | 2,555 | 141 | 4,390 |
| **Total** | **244,637** | **637,891** | **35,657** | **918,185** |

### Evaluation of T2D PRS performance

To identify an optimal T2D PRS for this investigation, in PAGE we assessed four distinct multi-ancestry PRS. The first three PRS are genome-wide PRS developed using 1.259 million HapMap3 variants in PRS-CSx^12^, an approach that has demonstrated high predictive ability across several traits^13–15^, and the fourth PRS includes 582 genome-wide significant T2D risk variants. **PRS Method 1**: a previously developed T2D PRS by Ge et al., 2022^16^ using meta-analyzed posterior effects with the META=T function in PRS-CSx and EUR GWAS summary statistics from Mahajan et al., 2018^17^, AFR GWAS summary statistics from the MEta-analysis of type 2 Diabetes in African Americans (MEDIA) Consortium^18^, and ASN GWAS summary statistics from Biobank Japan^19^; **PRS Method 2**: we developed a T2D PRS using a similar approach as PRS Method 1 but leveraging T2D GWAS summary statistics from a South Asian (SAS) population from Mahajan et al., 2022^6^ and from EUR, AFR, HIS, and ASN populations from Vujkovic et al., 2020^7^; **PRS Method 3**: we developed another T2D PRS calculated from a linear combination of population-specific PRS using the META=F function in PRS-CSx and the same summary statistics as PRS Method 2; and **PRS Method 4**: a previously developed multi-ancestry PRS with 582 genome-wide significant variants reported in Vujkovic et al., 2020^7,8^ (**Figure 1, Table S2, Methods**). PRS derived from each method were standardized to the distribution of controls within each population.

Each of the four T2D PRS methods exhibited good predictive performance in PAGE (**Table S3)**. Although confidence intervals of area under the receiver operating characteristic curve (AUC) overlapped between the methods, PRS Method 1 showed slightly better predictive performance across most populations compared to other methods. PRS Method 1 had AUC improvements of 1.8%, 4.0%, 5.2%, 4.8%, 4.5%, and 4.1% for AFR, AI, ASN, EUR, HIS, and NH populations, respectively, compared to the base model of age, sex, BMI, and the first 10 genetic principal components (PCs) (**Figure 2A and Table S3**). We used PRS Method 1 for all subsequent analyses.

**Figure 2.**
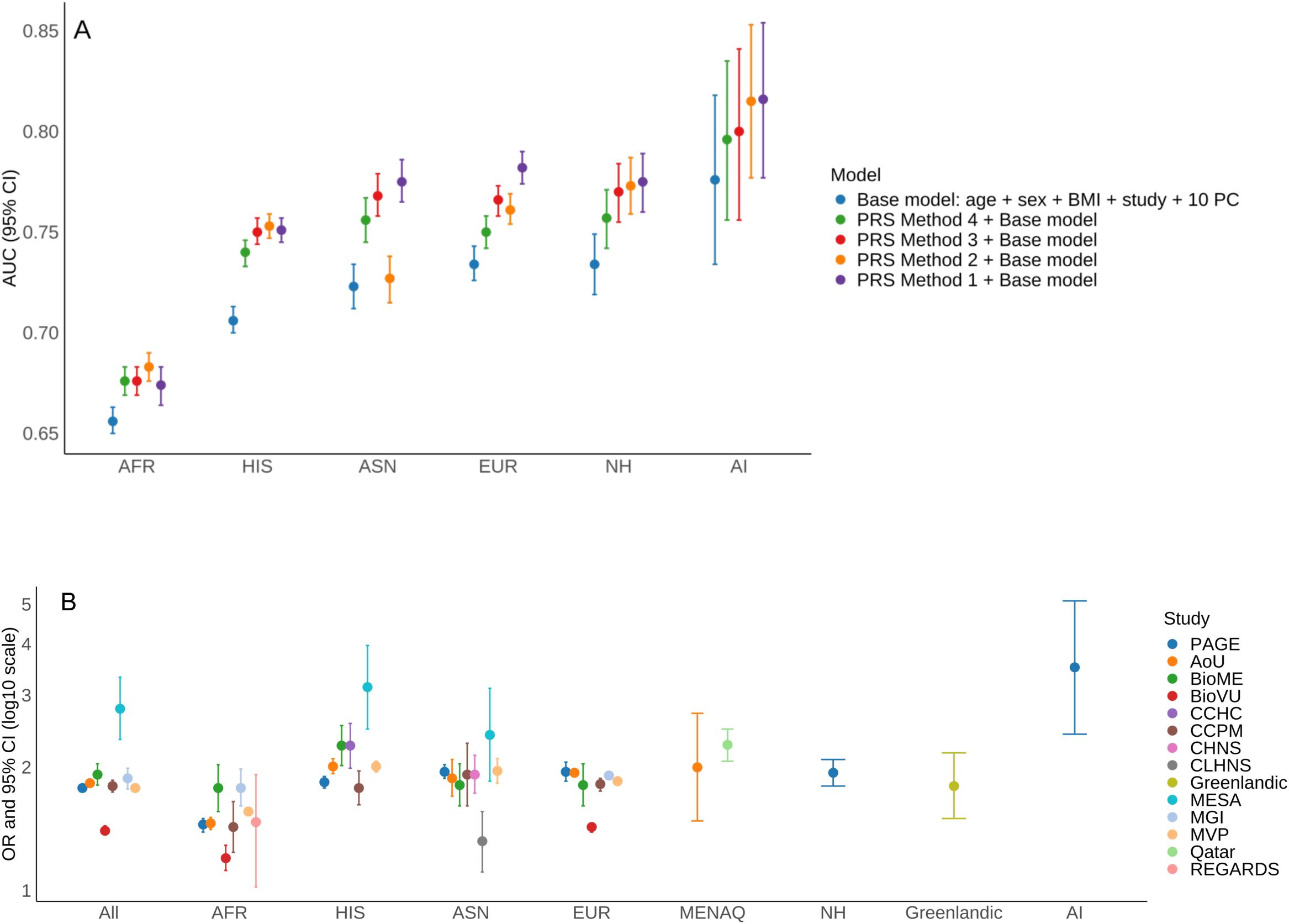
Performance of T2D PRS across self-identified populations. A) Area under the curve (AUC) for four T2D PRS methods in PAGE. Methods 1-4 correspond are defined in Figure 1. B) Association between each SD unit increase in the Ge et al. T2D PRS with T2D risk in PAGE and the additional biobanks and cohorts.

In PAGE, the PRS was highly predictive of T2D risk across populations but with variable performance (**Figure S1**). Each standard deviation (SD) unit increase in PRS was associated with 1.78- (95% CI: 1.75, 1.82) fold higher odds of T2D across all populations combined, ranging from 1.45- (95% CI: 1.39, 1.50) in AFR to 3.51- (95% CI: 2.41, 5.10) in AI (**Figure 2B and Table S4**). Compared to individuals in the 40-60% PRS category, the top 90-100% PRS decile was associated with 2.37 (95% CI: 2.24, 2.51) increased odds of T2D across all populations combined, ranging from 1.75 (95% CI: 1.54, 2.00) in AFR to 6.96 (95% CI: 2.25, 21.56) in AI (**Figure S2**). Effects were generally similar in the 13 additional biobanks and cohorts within each population (**Figure 2**).

### Context-dependent effects of T2D PRS

We investigated whether the T2D PRS demonstrated context-dependent effects on T2D risk by conducting association analyses stratified by 18 demographic, medical, and lifestyle and behavioral characteristics (**Methods and Table S5)**. Across PAGE and the additional biobanks and cohorts, we conducted stratified analyses in each of four populations (AFR, ASN, EUR, HIS) with sufficient sample sizes, as well as in all populations combined. We consistently found significant differences in T2D PRS effects by age groups, sex, family history (FH) of T2D, hypertension status, and obesity status within and across populations (**Figure 3 and Tables S6-S9**).

**Figure 3.**
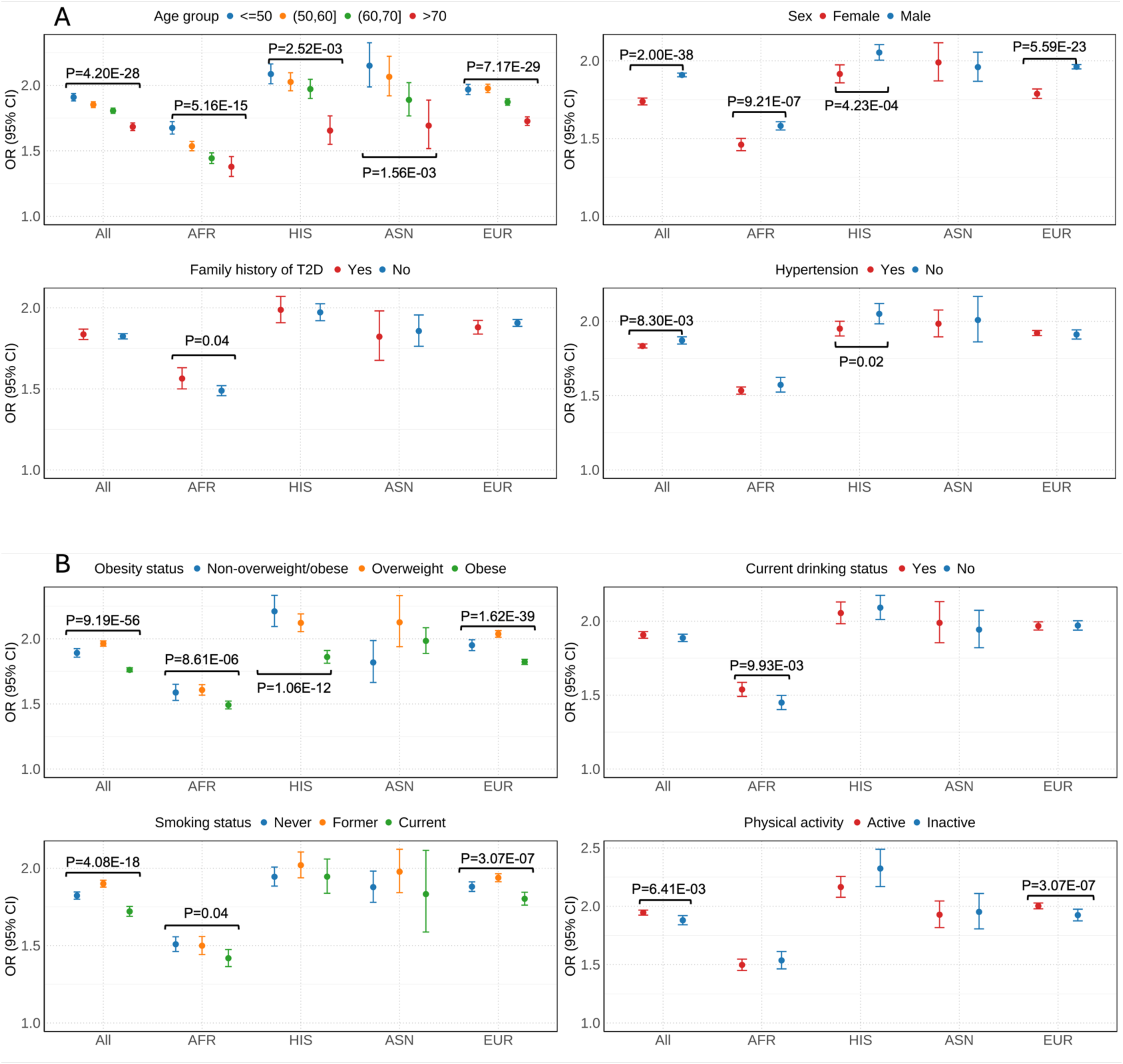
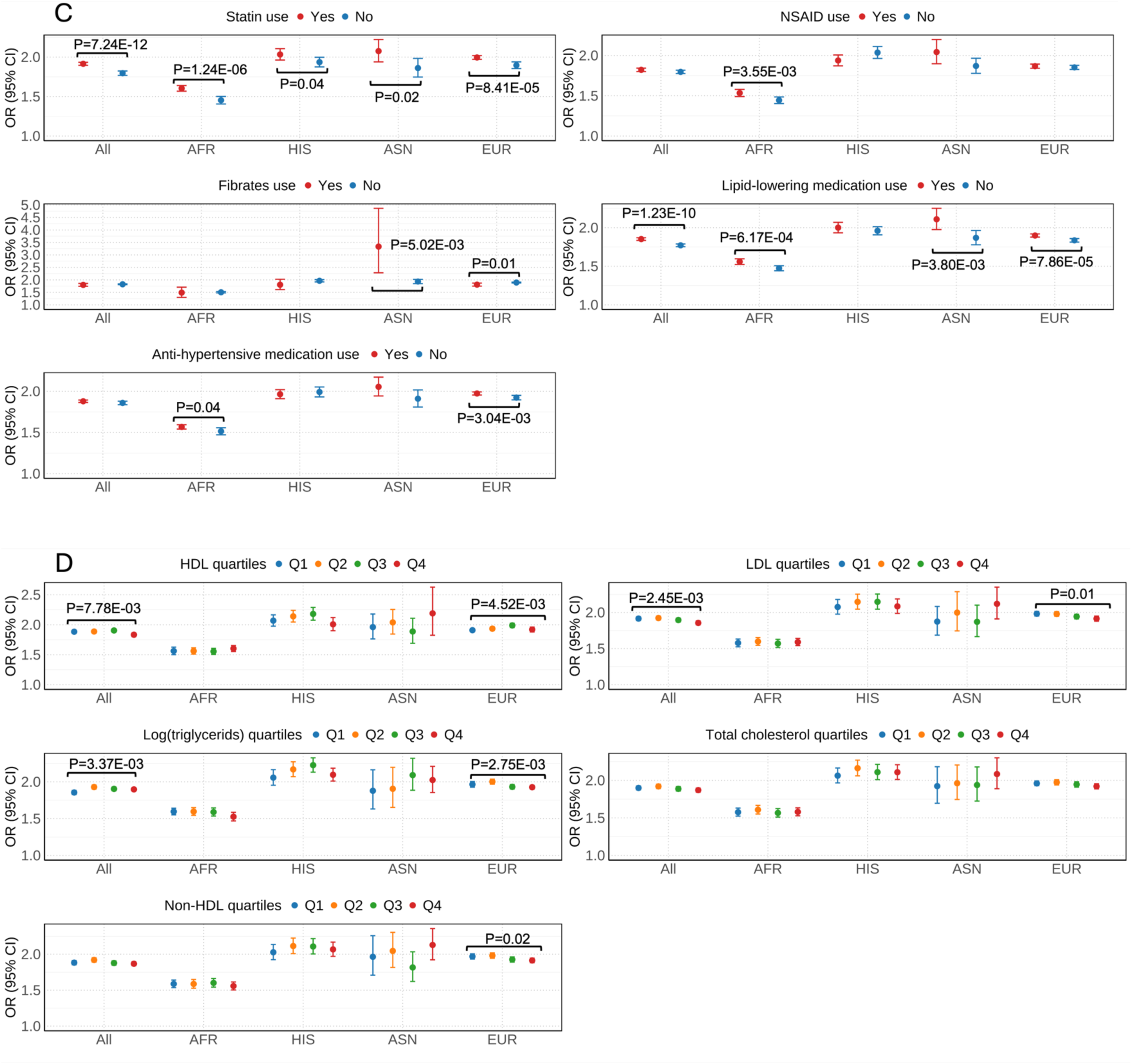
Effect of the T2D PRS on T2D risk stratified by demographic, medical, and lifestyle and behavioral factors meta-analyzed across PAGE and the additional biobanks and cohorts. P-values of heterogeneity are indicated when differences were statistically significant. A) Demographic characteristics and medical history factors. B) Behavioral and lifestyle factors. C) Medication use. D) Lipids.

Specifically, in meta-analyses across all populations, the T2D PRS was associated with significantly higher risk of T2D in younger individuals (age $50 years: OR=1.91 (95% CI: 1.88, 1.94); age 50-60 years: OR=1.85 (95% CI: 1.83, 1.88); age 60-70 years: OR=1.81 (95% CI: 1.79, 1.83); age >70 years: OR=1.68 (95% CI: 1.66, 1.71); P_het_=4.20×10^-28^). The T2D PRS also had a significantly larger effect in males compared to females (males: OR=1.91 (95% CI: 1.90, 1.92); females: OR=1.74 (95% CI: 1.72, 1.76); P_het_= 2.00×10^-38^). The effect of T2D PRS was slightly higher in individuals without hypertension (without hypertension: OR=1.87 (95% CI: 1.85, 1.90); with hypertension: OR=1.83 (95% CI: 1.82, 1.85); P_het_=8.30×10^-03^). The effect of the T2D PRS was attenuated in those with BMI categorized as obese (non-overweight/obese: OR=1.89 (95% CI: 1.86, 1.92); overweight: OR=1.96 (95% CI: 1.94, 1.98); obese: OR=1.76 (95% CI: 1.75, 1.78); P_het_ =9.19×10^-56^). Although no significant PRS performance difference by FH of T2D was observed across all populations, a stronger effect was observed among those with a FH of T2D in AFR populations (FH: OR=1.56 (95% CI: 1.50, 1.63); without FH: OR=1.49 (95% CI: 1.46, 1.52); P_het_=0.04). When separately investigating PAGE versus the additional biobanks and studies, similar trends across age groups, sex, hypertension and obesity status were observed (**Figure S3 and Tables S6, S8, S9**).

PRS had a stronger effect in never smokers and former smokers compared to current smokers (P_het_=4.08×10^-18^) and in physically active individuals to inactive individuals (P_het_ =6.14×10^-03^) in meta-analyses across all populations, although these findings were not consistent when examining each population separately. Additionally, the PRS effect was stronger in statin and lipid-lowering medication users than non-users, though the differential association is complicated by higher statin use among diabetes cases and incomplete documentation of lipid medication use. Findings were inconsistent for other medication use, lipids, and current drinking status.

### Association between T2D PRS and diabetes-related traits

Next, to investigate the clinical implications of genetic risk of T2D, we examined associations between the T2D PRS and 20 diabetes-related traits across five primary phenotype categories: glycemic traits, cardiometabolic traits, vascular diseases, kidney diseases and functions, and inflammatory biomarkers, stratified by T2D status in AFR, ASN, EUR, HIS, and all populations combined (**Methods and Table S10**). Significant associations were found across various glycemic and cardiometabolic traits, including HbA1c, fasting glucose, hypertension, systolic blood pressure (SBP), diastolic blood pressure (DBP), triglycerides, non-HDL, and HDL at a *P* < 2.50 × 10^-3^ (0.05/20 traits) Bonferroni adjusted threshold (**Figure 4 and S4, Tables S11-**S13).

**Figure 4.**
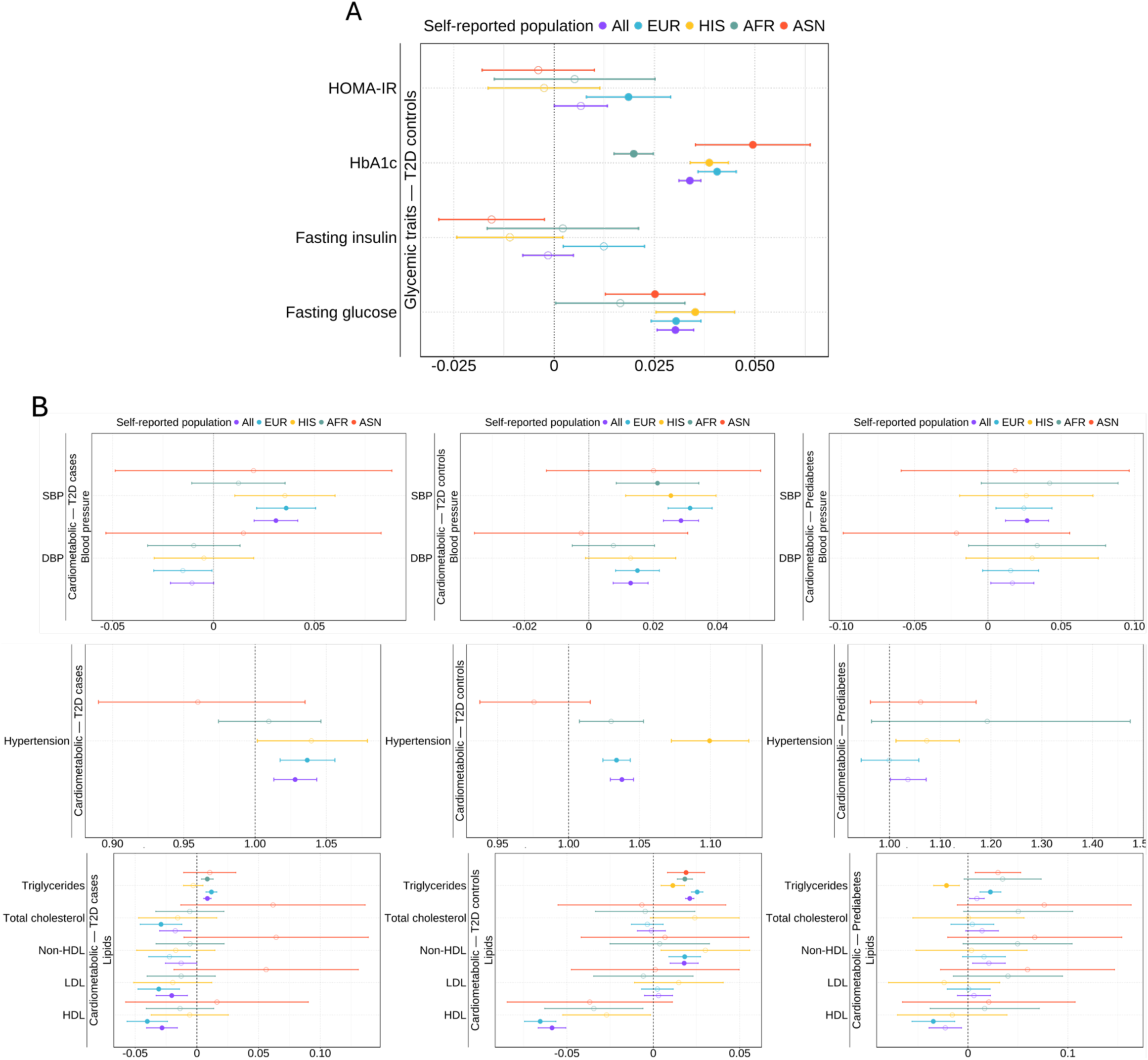
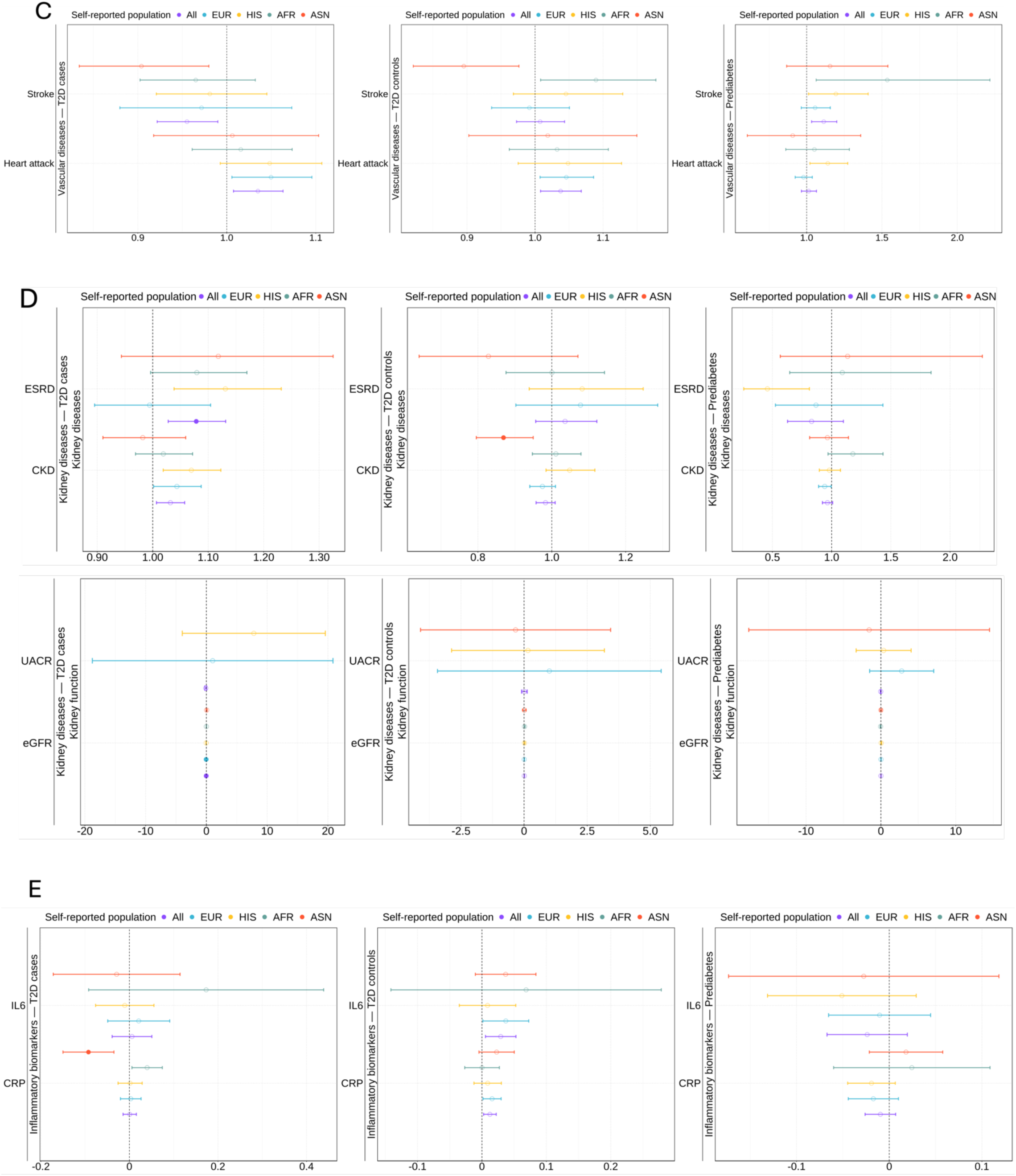
Effect of T2D PRS on diabetes-related traits meta-analyzed across PAGE and the additional biobanks and cohorts. The X-axis represents the beta estimates and 95% CIs of PRS for continuous traits and odds ratios and 95% CIs of PRS for binary traits. Solid circles indicate significant associations that passed the Bonferroni-adjusted P-value threshold of *P* < 2.50 × 10^−3^, while open circles indicate associations that were not significant. Association between T2D PRS and A) glycemic traits in T2D controls. B) cardiometabolic traits in T2D cases, controls, and individuals with prediabetes. C) vascular diseases in T2D cases, controls, and individuals with prediabetes. D) kidney function related traits in T2D cases, controls, and individuals with prediabetes. E) inflammatory biomarkers in T2D cases, controls, and individuals with prediabetes. The units of continuous traits are listed in **Table S10**.

We first examined the associations between T2D PRS and glycemic traits among T2D controls, as glycemic measurements in T2D cases are influenced by diabetes medications. Each SD increase in the T2D PRS was significantly associated with 0.034 (95% CI: 0.031, 0.037) higher units of HbA1c levels and 0.030 mmol/L (95% CI: 0.026, 0.035) higher fasting glucose levels in individuals without T2D (**Figures 4A**). These findings were consistent across population and studies. Additionally, the PRS demonstrated a significant association with log transformed HOMA-IR (P=5.19×10^-04^), although this was observed only in the EUR population.

Regarding cardiometabolic traits, each SD increase in PRS was associated with a 1.03-fold increased risk (95% CI: 1.03, 1.05) of hypertension in T2D controls across all populations, a 1.04-fold (95% CI: 1.02, 1.06) and 1.03-fold (95% CI: 1.02, 1.04) increased risk in EUR cases and controls, and a 1.10-fold (95% CI: 1.07, 1.13) increased risk in HIS controls (**Figures 4B**). PRS was also positively associated with SBP (*P*=8.24E-25), DBP (*P*=2.72E-06), log-transformed triglycerides (*P*=1.13E-68), and non-HDL (*P*=1.50E-05), and negatively associated with HDL (*P*=1.50E-05) in T2D controls across all populations.

For kidney diseases, the PRS was associated with a 1.08-fold increased risk (95% CI: 1.03, 1.13) of end stage renal disease (ESRD) in T2D cases across all populations. However, PRS was inversely associated with a 0.87-fold increased risk (95% CI: 0.80, 0.95) of chronic kidney disease (CKD) in ASN controls. The PRS was not consistently associated with any vascular diseases or inflammatory markers.

### T2D PRS PheWAS

To further investigate the prognostic value of the T2D PRS and clinical phenotypes, we conducted a comprehensive PheWAS meta-analysis of the T2D PRS on 1,815 unique phenotypes representing 17 disease categories tested across five biobanks. We identified significant associations between the T2D PRS and 732 traits (40.3% of all phecodes tested) across all populations combined at Bonferroni adjusted significance (*P* < 2.75 × 10^-5^), as well as 213 (12.0%), 43 (3.7%), 689 (38.0%), and 232 (13.7%) significant traits in AFR, ASN, EUR, and HIS populations, respectively (**Figures 5, S4, and Tables 1, S14**). All significant traits identified in AFR, ASN, and HIS and were also significant in the EUR population, which had an additional 428 significant traits not found in any other population (**Figure 6**). The PRS-PheWAS effect estimates (ORs) in EUR were reasonably correlated with AFR (r=0.68), ASN (r=0.64), and HIS (r=0.73) populations, with consistent effect directions. Effects had slightly lower correlations between AFR and HIS (r=0.58), ASN and AFR (0.58), and ASN and HIS (0.59), likely reflecting the larger sample size in the EUR population, which allowed more accurate estimates of effects and thus more overlap with other populations.

**Figure 5.**
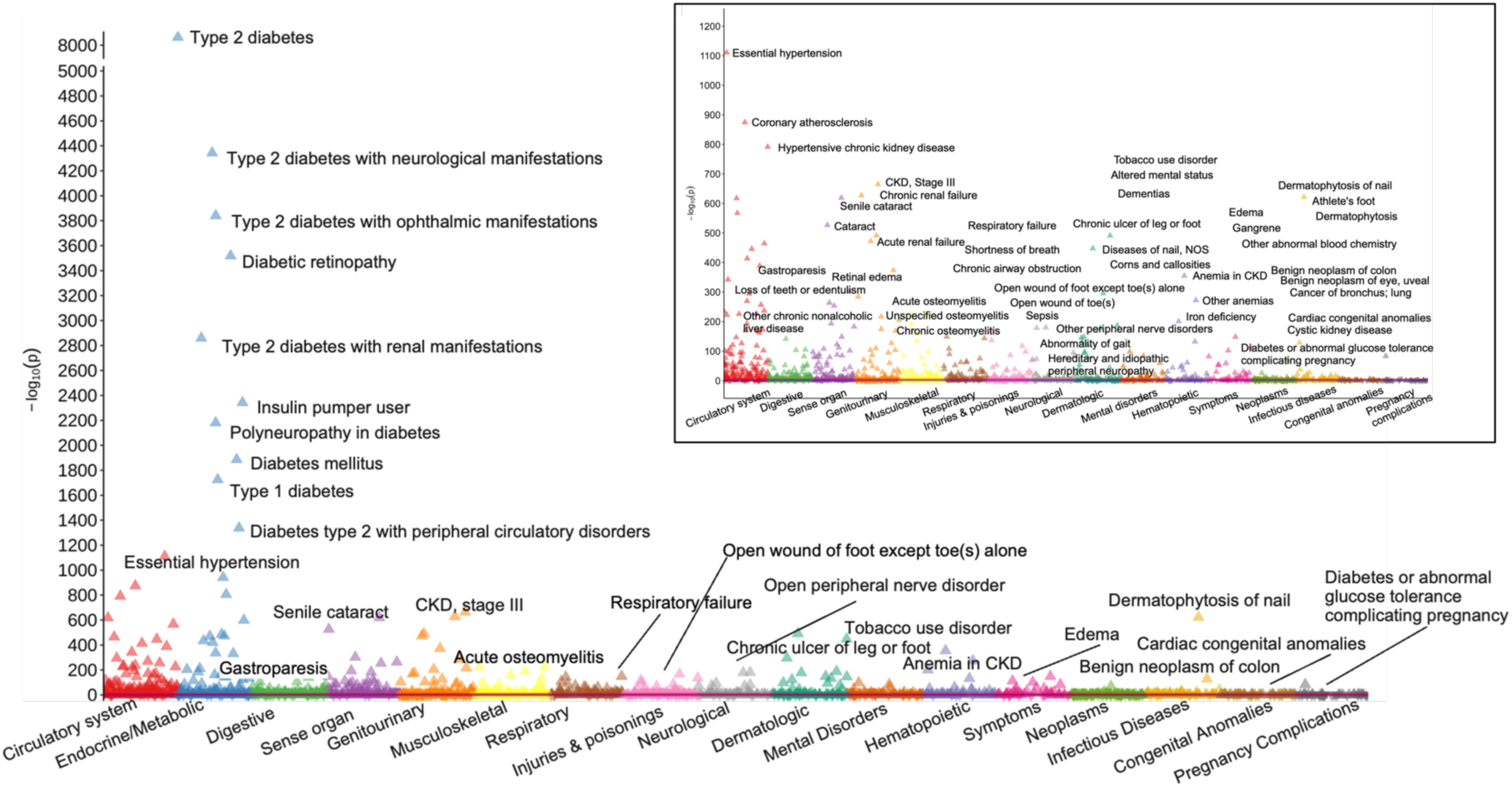
T2D PRS PheWAS results meta-analyzed across all five biobanks and populations. The X-axis represents phecodes color-coded by their corresponding phenotype category and is ordered by the category with the most to the category with the least significant hits. The Y-axis represents the −log10(p-value). The red horizontal line represents Bonferroni-adjusted p-value threshold *P* < 2.75 × 10^−5^ (732 phecodes meet this threshold), and the blue horizontal line represents an unadjusted p-value threshold of *P* < 0.05 (1,120 phecodes meet this threshold). Upward triangles indicate positive associations, while downward triangles indicate negative associations. The top ten most significant associations from the endocrine/metabolic category are annotated, while the single most significant association from each other category is annotated. The inset plot excludes the endocrine/metabolic category and annotates the top three PheWAS result in each category.

**Figure 6.**
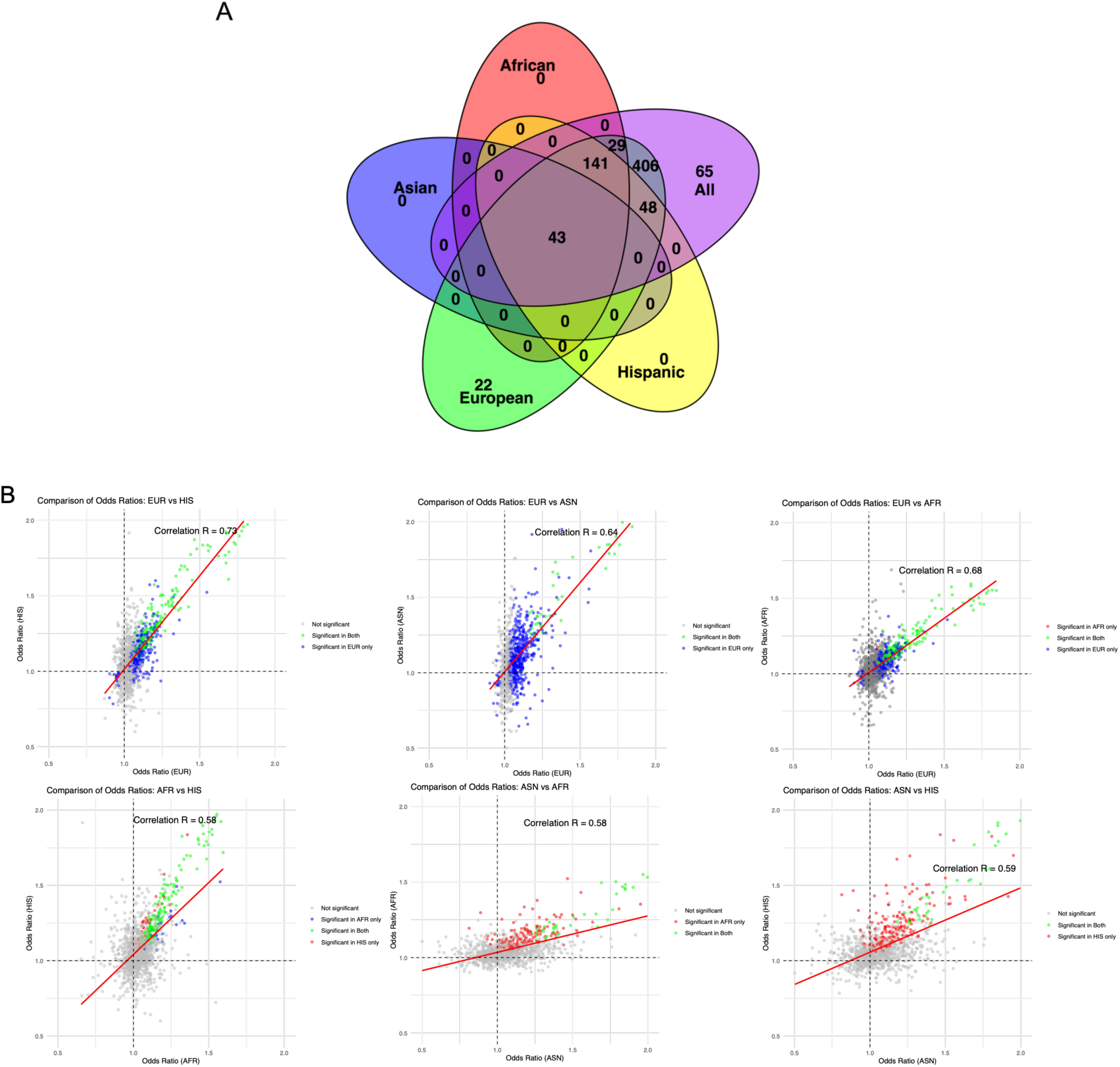
Comparison of significant associations and effect sizes of T2D PRS PheWAS results by population. A) Venn diagram of significant PheWAS results across populations. B) Comparison of odds ratios from T2D PRS PheWAS results across populations.

The 732 significant associations encompassed phenotypes related to the circulatory system (n=123, 16.8%), endocrine and metabolic functions (n=90, 12.3%), digestive system conditions (n=82, 11.2%), sense organs (n=65, 8.9%), genitourinary system conditions (n=58, 7.9%), and 12 other disease categories (n=314, 42.9%). In analyses conducted across all populations combined, T2D (OR=2.38; *P*-value<5.0E-324) demonstrated the most significant association, followed by T2D related phenotypes from the endocrine/metabolic phenotype group, including diabetic retinopathy, insulin pump user, polyneuropathy in diabetes, and diabetes mellitus (**Table 2**, **Figures 5 and S14**). We observed consistently strong associations overall and across populations between the T2D PRS and type 1 diabetes (T1D) risk, with each SD increase in the T2D PRS associated with 1.63-(*P*=1.31E-173), 1.47-(*P*=1.31E-173), 1.83-(*P*=2.97×10^-18^), 1.63-(*P*<5.0E-324), and 1.76-(*P*=4.41E-128), and fold increased risk of T1D in the overall, AFR, ASN, EUR, and HIS populations, respectively.

**Table 2.** Significant meta-analyzed phenome-wide associations (meta-PheWAS) between T2D PRS and clinical diseases across all populations. If a Phecode with the same one-decimal value (e.g., 250.2) is listed, more specific phecodes (e.g., 250.24) are excluded; otherwise, the specific phecode is shown. The top 10 associations in the endocrine/metabolic disease category are shown, while the top 5 associations are shown for other phenotype categories. All associations shown passed Bonferroni-adjusted p-value threshold *P* < 2.75 × 10^-5^. Full meta-PheWAS results are provided in Table S14.

| **Phecode** | **Phenotype** | **Group** | **OR** | **P-value** |
| --- | --- | --- | --- | --- |
| 250.2 | Type 2 diabetes | Endocrine/Metabolic | 2.38 | <5E-324 |
| 250.7 | Diabetic retinopathy | Endocrine/Metabolic | 2.52 | <5E-324 |
| 250.3 | Insulin pump user | Endocrine/Metabolic | 2.19 | <5E-324 |
| 250.6 | Polyneuropathy in diabetes | Endocrine/Metabolic | 2.23 | <5E-324 |
| 250 | Diabetes mellitus | Endocrine/Metabolic | 1.80 | <5E-324 |
| 250.1 | Type 1 diabetes | Endocrine/Metabolic | 2.05 | <5E-324 |
| 272.1 | Hyperlipidemia | Endocrine/Metabolic | 1.27 | <5E-324 |
| 249 | Secondary diabetes mellitus | Endocrine/Metabolic | 2.05 | <5E-324 |
| 250.4 | Abnormal glucose | Endocrine/Metabolic | 1.36 | <5E-324 |
| 276.13 | Hyperpotassemia | Endocrine/Metabolic | 1.36 | <5E-324 |
| 401.1 | Essential hypertension | Circulatory System | 1.34 | <5E-324 |
| 411.4 | Coronary atherosclerosis | Circulatory System | 1.33 | <5E-324 |
| 401.22 | Hypertensive chronic kidney disease | Circulatory System | 1.48 | <5E-324 |
| 411.8 | Other chronic ischemic heart disease, unspecified | Circulatory System | 1.38 | <5E-324 |
| 443.9 | Peripheral vascular disease, unspecified | Circulatory System | 1.40 | <5E-324 |
| 536.3 | Gastroparesis | Digestive | 1.46 | 6.66E-142 |
| 525.1 | Loss of teeth or edentulism | Digestive | 1.16 | 4.55E-106 |
| 571.5 | Other chronic nonalcoholic liver disease | Digestive | 1.14 | 4.42E-104 |
| 530.11 | GERD | Digestive | 1.07 | 9.83E-83 |
| 525 | Other diseases of the teeth and supporting structures | Digestive | 1.12 | 7.09E-69 |
| 366.2 | Senile cataract | Sense Organs | 1.27 | <5E-324 |
| 366 | Cataract | Sense Organs | 1.27 | <5E-324 |
| 362.9 | Retinal edema | Sense Organs | 1.72 | 3.90E-303 |
| 367 | Disorders of refraction and accommodation; blindness and low vision | Sense Organs | 1.18 | 2.61E-264 |
| 362.3 | Other nondiabetic retinopathy | Sense Organs | 1.40 | 2.01E-254 |
| 585.3 | Chronic renal failure | Genitourinary | 1.34 | <5E-324 |
| 585.1 | Acute renal failure | Genitourinary | 1.28 | <5E-324 |
| 588.2 | Secondary hyperparathyroidism (of renal origin) | Genitourinary | 1.52 | 1.69E-217 |
| 585.2 | Renal failure NOS | Genitourinary | 1.44 | 2.50E-206 |
| 585.4 | Chronic kidney disease, Stage I or II | Genitourinary | 1.29 | 1.99E-169 |
| 710.11 | Acute osteomyelitis | Musculoskeletal | 1.65 | 1.55E-225 |
| 710.19 | Unspecified osteomyelitis | Musculoskeletal | 1.49 | 1.91E-224 |
| 710.12 | Chronic osteomyelitis | Musculoskeletal | 1.58 | 9.49E-192 |
| 735.21 | Hammer toe (acquired) | Musculoskeletal | 1.26 | 9.47E-161 |
| 741.3 | Difficulty in walking | Musculoskeletal | 1.20 | 1.81E-135 |
| 509.1 | Respiratory failure | Respiratory | 1.19 | 1.44E-149 |
| 512.7 | Shortness of breath | Respiratory | 1.11 | 3.05E-144 |
| 496 | Chronic airway obstruction | Respiratory | 1.12 | 4.49E-109 |
| 512.9 | Other dyspnea | Respiratory | 1.09 | 1.85E-90 |
| 480 | Pneumonia | Respiratory | 1.12 | 7.58E-76 |
| 871.3 | Open wound of foot except toe(s) alone | Injuries & Poisonings | 1.49 | 2.97E-168 |
| 871.4 | Open wound of toe(s) | Injuries & Poisonings | 1.58 | 3.28E-137 |
| 994.2 | Sepsis | Injuries & Poisonings | 1.19 | 4.84E-118 |
| 854 | Complications of cardiac/vascular device, implant, and graft | Injuries & Poisonings | 1.32 | 1.39E-98 |
| 872 | Traumatic amputation | Injuries & Poisonings | 1.49 | 1.49E-86 |
| 351 | Other peripheral nerve disorders | Neurological | 1.16 | 5.25E-181 |
| 350.2 | Abnormality of gait | Neurological | 1.15 | 1.56E-179 |
| 356 | Hereditary and idiopathic peripheral neuropathy | Neurological | 1.17 | 3.67E-96 |
| 337.1 | Peripheral autonomic neuropathy | Neurological | 1.23 | 1.36E-83 |
| 327.3 | Sleep apnea | Neurological | 1.08 | 1.36E-77 |
| 707.2 | Chronic ulcer of leg or foot | Dermatologic | 1.54 | <5E-324 |
| 703 | Diseases of nail, NOS | Dermatologic | 1.44 | <5E-324 |
| 700 | Corns and callosities | Dermatologic | 1.38 | 1.46E-296 |
| 681.1 | Cellulitis and abscess of fingers/toes | Dermatologic | 1.30 | 2.68E-188 |
| 681.6 | Cellulitis and abscess of foot, toe | Dermatologic | 1.55 | 2.12E-178 |
| 318 | Tobacco use disorder | Mental Disorders | 1.09 | 1.07E-98 |
| 292.4 | Altered mental status | Mental Disorders | 1.15 | 3.76E-85 |
| 290.1 | Dementias | Mental Disorders | 1.18 | 1.86E-60 |
| 290 | Delirium dementia and amnestic and other cognitive disorders | Mental Disorders | 1.13 | 1.78E-30 |
| 317.1 | Alcoholism | Mental Disorders | 0.94 | 1.17E-27 |
| 285.21 | Anemia in chronic kidney disease | Hematopoietic | 1.50 | <5E-324 |
| 285 | Other anemias | Hematopoietic | 1.17 | 3.36E-273 |
| 280.1 | Iron deficiency anemias, unspecified or not due to blood loss | Hematopoietic | 1.19 | 1.64E-201 |
| 286.2 | Encounter for long-term (current) use of anticoagulants | Hematopoietic | 1.11 | 1.25E-61 |
| 285.1 | Acute posthemorrhagic anemia | Hematopoietic | 1.13 | 6.90E-52 |
| 782.3 | Edema | Symptoms | 1.15 | 2.91E-149 |
| 791 | Gangrene | Symptoms | 1.75 | 9.85E-114 |
| 790.6 | Other abnormal blood chemistry | Symptoms | 1.13 | 9.12E-103 |
| 772.3 | Muscle weakness | Symptoms | 1.15 | 5.00E-83 |
| 789 | Nausea and vomiting | Symptoms | 1.09 | 1.62E-59 |
| 208 | Benign neoplasm of colon | Neoplasms | 1.08 | 7.55E-70 |
| 224.1 | Benign neoplasm of eye, uveal | Neoplasms | 1.12 | 2.03E-26 |
| 165.1 | Cancer of bronchus; lung | Neoplasms | 1.08 | 7.90E-13 |
| 172 | Skin cancer | Neoplasms | 0.95 | 4.39E-12 |
| 155.1 | Malignant neoplasm of liver, primary | Neoplasms | 1.14 | 8.56E-12 |
| 110.1 | Dermatophytosis | Infectious Diseases | 1.10 | 1.38E-38 |
| 38 | Septicemia | Infectious Diseases | 1.11 | 3.94E-28 |
| 110 | Dermatophytosis / Dermatomycosis | Infectious Diseases | 1.10 | 6.17E-23 |
| 41 | Bacterial infection NOS | Infectious Diseases | 1.08 | 9.33E-23 |
| 41.1 | Staphylococcus infections | Infectious Diseases | 1.14 | 4.21E-20 |
| 747.1 | Cardiac congenital anomalies | Congenital Anomalies | 1.06 | 1.45E-07 |
| 751.21 | Cystic kidney disease | Congenital Anomalies | 1.06 | 2.51E-07 |
| 649.1 | Diabetes or abnormal glucose tolerance complicating pregnancy | Pregnancy Complications | 1.46 | 6.32E-84 |

The single top phenotype associated with the T2D PRS from other phenotype groups, all of which were positively associated, were essential hypertension, CKD, senile cataract, chronic ulcer of leg or foot, anemia in chronic kidney disease, acute osteomyelitis, other peripheral nerve disorders, open wound of foot, respiratory failure, edema, gastroparesis, tobacco use disorder, diabetes or abnormal glucose tolerance complicating pregnancy disorder, benign neoplasm of colon, dermatophytosis, and cardiac congenital anomalies.

## Discussion

In this large-scale T2D investigation of more than 240,000 T2D cases and 630,000 controls from PAGE and 13 independent biobanks and studies, we comprehensively characterized a multi-ancestry genome-wide T2D PRS across diverse populations. We conducted a harmonized assessment of PRS performance to evaluate its context-dependent effects across different demographic and clinical characteristics. Findings from this investigation demonstrate that the PRS holds predictive value for T2D risk across diverse populations. The T2D PRS demonstrated notable context-dependent associations with T2D risk, with PRS performance varying by certain demographic and clinical characteristics, such as age, sex, FH of T2D, hypertension, and obesity status. Additionally, we found that genetic risk of T2D, as measured by the T2D PRS, was associated with other glycemic and cardiometabolic traits in controls, suggesting that PRS can serve as a valuable tool for predicting dysglycemia and metabolic risk, thereby facilitating early interventions and improving clinical risk assessment. PheWAS results found associations between the T2D PRS and 40.3% of tested phenotypes, including phenotypes across all disease categories tested. These findings have notable implications for the prognostic value and clinical utility of T2D PRS.

We found that a multi-ancestry PRS was strongly associated with increased risk of T2D in all populations examined, yielding an overall OR of 1.75 (95% CI: 1.71, 1.78) and AUC of 0.641 (95% CI: 0.636, 0.645). The PRS performance across studies was generally comparable, with small variations likely due to study-specific differences, such as study design and phenotype definition. However, we observed larger variations in the performance of T2D PRS across populations, with the highest predictive performance in the AI population (although sample sizes were small), followed by EUR, ASN, HIS, and AFR populations. This performance variability could be attributed to various population-specific factors that are likely not strongly represented in the discovery GWAS used to construct the T2D PRS.^20^ This notably underscores the importance of expanding GWAS and fine-mapping efforts, particularly in AFR populations, to improve the accuracy and transferability of T2D PRS.

Our results show that that prediction accuracy of PRS is context-dependent and can vary even within populations. We consistently observed that the T2D PRS was particularly informative for predicting T2D susceptibility in younger individuals, those with a FH of T2D, those without hypertension, males, and individuals who were not obese or overweight, across and within populations. The stronger predictive ability in younger individuals is consistent with published literature, such as the findings from the Framingham Offspring Study^21^ and the EPIC InterAct Study^9^, both of which observed a higher relative effect of T2D PRS among individuals under 50 years old compared to those over 50. However, these studies have been limited to T2D PRS developed and evaluated in individuals predominantly of European descent. Similar trends have also been observed in other traits, including prostate cancer^22–24^, colorectal cancer^25,26^, estrogen receptor-positive breast cancer^27^, and coronary artery disease^28^. The stronger genetic influence on disease onset at a younger age is expected, as older individuals have had more time to accumulate environmental risk factors that lead to metabolic impairments, thereby impacting disease risk. The larger estimated effect of T2D PRS in leaner adults and those without hypertension observed in this study could also be attributed to the higher risk contribution of genetic factors in those with a lower environmental contribution to T2D, a pattern also shown in several previous studies^8,9,29^. Additionally, the larger effect size of the PRS in individuals with a FH suggests potential interactions of the PRS with rare genetic variants not included in the PRS but shared within families, or interactions of the PRS with environmental risk factors shared within families. This finding highlights partial independence of PRS and FH in predicting disease risk, suggesting that polygenic risk for T2D and FH are not redundant disease risk factors.

It also emphasizes the importance of integrating FH information with PRS for more accurate risk stratification and prediction, as demonstrated for other traits^30,31^. Prior research on the performance of T2D PRS stratified by sex is limited^9,32^, and evidence of sex-based differences in T2D heritability estimates is inconsistent^33–36^. However, sex-based differences in PRS prediction have been widely reported in other traits.^37–39^ The sex-based differences observed in our study could be attributed to several factors, including alleles exhibiting sex differential effects for T2D^40,41^, diagnosis bias leading to T2D being more frequently diagnosed in males^42^, differences in exposure to environmental and lifestyle factors that affect T2D risk between males and females, or phenotypic differences, such as hormones and body composition^40,43^. Future sex-specific GWAS and gene-by-sex interaction studies are needed to further investigate sex-specific genetic factors in T2D. Collectively, these interactions between the T2D PRS and non-genetic factors demonstrate the complex nature of T2D risk and the importance of interpreting genetic risk of T2D in the context of non-genetic risk factors. This indicates that risk prediction models will need to include not only PRS but demographic, medical, lifestyle and behavioral characteristics, and their interactions.^37^ Moreover, any potential application of PRS in clinical care will need to consider such contextual factors.

Our study provides compelling evidence supporting the prognostic value of T2D PRS for various diabetes-related traits and disease conditions across diverse populations. Our association analyses of 20 diabetes-related traits showed significant associations between T2D PRS and glycemic parameters, such as fasting glucose and HbA1c, in T2D controls, as well as cardiometabolic traits, including hypertension, SBP, triglycerides, and HDL, in both T2D cases and controls. These findings suggest that T2D PRS may serve as a valuable tool for early identification of individuals at risk for developing dysglycemia and cardiometabolic complications, even among those who are currently asymptomatic or undiagnosed. The inverse association we observed between T2D PRS and CKD in ASN controls could be due to selection bias, where individuals with higher genetic risk for T2D who remain diabetes-free may represent a healthier subset of participants, possibly benefiting from protective environmental or lifestyle factors that also lower their risk of CKD. Future research is needed to validate this finding.

Findings from our PheWAS across 1,815 diseases and conditions and five biobanks further demonstrated significant associations between T2D PRS and various clinical comorbidities, including those related to circulatory system, endocrine and metabolic disorders, lipid abnormalities, and kidney diseases. Notably, these associations were consistently observed across populations, with fairly correlated effect estimates. Despite the variability in PRS performance for T2D risk estimation across populations, our findings suggest that genetic mechanisms underlying T2D and diabetes-related traits are shared across populations. As such, the PRS may serve as a valuable tool for stratifying the risk of complications in individuals diagnosed with T2D across diverse populations. The positive association between T2D PRS and micro- and macrovascular complications, such as retinopathy, neuropathy, CKD, cardiovascular diseases, and peripheral artery disease have also been reported by prior studies.^7,44,45^ The significant positive associations between the T2D PRS and T1D risk from PheWAS may indicate the potential misclassification of T2D and T1D in clinician-recorded diagnosis, given the overlapping clinical features T2D and T1D^46^, or it could reflect genetic consistencies between two diseases. This study complements prior genetic research on T2D by leveraging large sample sizes to identify significant associations between T2D PRS and various traits and conditions not previously reported. Our findings imply that the T2D PRS holds promise as a powerful tool for precision medicine, risk stratification, and disease management in the context of T2D, providing an opportunity to identify individuals at elevated risk of developing T2D and its complications.

Further, the PheWAS results reported here further serve as a comprehensive catalog of phenotypic associations with T2D PRS, contributing to our understanding of T2D-related comorbidity patterns. These findings could offer a valuable resource for hypothesis generation, disease subtype characterization, and the identification of shared biological pathways and molecular mechanisms.

The results of our study should be interpreted considering several limitations. We used self-identified race and ethnicity (SIRE) population descriptors to determine whether the T2D PRS had discrepancies in performance in the contexts of SIRE. Although other sociodemographic and environmental factors may be more informative than SIRE, because our investigation included a large number of studies, many of which include legacy data collected up to 37 years ago, this study was limited by the variables available across all included studies. Further, SIRE information was collected differently between studies (**Table S14**), which could influence PRS performance between populations. In addition, this study focused on prevalent T2D. Future research should replicate these findings by investigating the association between PRS and risk of incident T2D and the other outcomes reported here. Further, most phenotypes and clinical measurements, such as glucose levels and BMI, were collected at baseline, when these measurements are typically available for most individuals. This limits our ability to capture changes over time and assess the dynamic nature of these associations.

In conclusion, our study highlights the predictive value of the T2D PRS across diverse populations, emphasizing its potential as a promising tool for identifying individuals at elevated risk of developing T2D and its associated comorbidities and complications, shedding light on potential avenues for risk prediction and personalized management strategies. Our findings also demonstrate that the T2D PRS has context dependent effects, indicating the need to account for these characteristics to interpret PRS and improve PRS prediction accuracy. Further research is warranted to evaluate and optimize the clinical utility of T2D PRS across diverse populations.

## Methods

### Participants

The PAGE Study was developed to conduct genetic epidemiological research in ancestrally diverse populations within the United States.^47^ PAGE includes individuals from the following ongoing population-based biobanks and cohorts: Atherosclerosis Risk in Communities (ARIC)^48^, the Icahn School of Medicine at Mount Sinai BioMe biobank in New York City (BioMe), Coronary Artery Risk Development in Young Adults Study (CARDIA)^49^, Hispanic Community Health Study/Study of Latinos (HCHS/SOL)^50,51^, Multiethnic Cohort Study (MEC)^52^, and Women’s Health Initiative (WHI)^53^. All participants provided informed consent, and each study was approved by the Institutional Review Board. A detailed description of PAGE has been described,^47^ with additional detail summarized in the **Supplemental Information** and **Table S1**.

Analyses were also conducted in 13 additional biobanks and cohorts that were not included in the discovery GWAS summary statistics of the best performing T2D PRS. Studies included All of Us (AoU)^54^, BioMe (participants were non-overlapping with those from BioMe included in PAGE), BioVU Biobank (BioVU)^55^, Cameron County Hispanic Study (CCHC)^56^, Cebu Longitudinal Health and Nutrition Survey (CLHNS)^57^, China Health and Nutrition Survey (CHNS)^58^, Colorado Center for Personalized Medicine Biobank (CCPM)^59^, Greenlandic Health Surveys (Greenlandic)^60–62^, Multi-Ethnic Study of Atherosclerosis Study (MESA)^63^, Michigan Genomics Initiative (MGI)^64^, Million Veteran Program (MVP)^65–67^, Qatar Biobank (Qatar)^68^, and REasons for Geographic and Racial Differences in Stroke Study (REGARDS)^69^ (**Table S1**). Analyses performed in these additional studies were dependent on the availability of variables and data needed to derive variables.

### Use of population descriptors

Self-identified race and ethnicity (SIRE) population descriptors were used across studies, with the goal of studying how PRS performance may vary across the contexts of SIRE. Although other sociodemographic and environmental factors are expected to be more informative than SIRE, this investigation is limited by the variables that are currently available across all included studies. It is important to note that SIRE does not serve as a proxy for genetic ancestry, holds no biological basis, and does not imply a biological explanation for health disparities.^70^

We standardized population descriptors across all studies in the analysis by using consistent acronyms. Specifically, AFR refers to African, African American, or Black participants; AI refers to American Indian, Alaskan Native, or Native American; ASN includes Asian American, East or Southeast Asian (e.g., from China, Japan, Korea, Indonesia, the Philippines), as well as South Asian/Indian (e.g., from India, Pakistan); EUR refers to non-Hispanic White American or European individuals; Greenlandic refers to individuals of Greenlandic or mixed Greenlandic-Danish heritage; HIS represents Hispanic/Latino American participants; MENAQ encompasses Qatari and Arab American individuals; and NH refers to Native Hawaiian or Other Pacific Islander. The Greenlandic population was considered separate from others as this population represents substantial Inuit ancestry.^61^ Detailed definitions of population descriptors used in each study are provided in **Table S15**.

### T2D and prediabetes definition

T2D and prediabetes definitions were created based on the American Diabetes Association’s 2021 Standards of Medical Care in Diabetes.^71^ In PAGE, participants were classified as T2D cases if they met at least one of the following criteria: adults ≥18 years old with 1) a T2D diagnosis by a physician/medical professional or use of medication for treatment of diabetes, 2) a fasting (≥8 hour) plasma glucose ≥126 mg/dl, 3) a random glucose ≥200 mg/dl, or 4) an HbA1c ≥48 mmol/mol (6.5%). Cases were restricted to those with either an age at diagnosis or most recent age at glucose or HbA1c draw ≥25 years to avoid misclassifying T1D cases as T2D cases. Individuals with prediabetes were defined as adults ≥18 years old who did not meet the definition of T2D and with 1) a fasting plasma glucose between 100-125 mg/dl, 2) an HbA1c between 39-46 mmol/mol (5.7–6.4%), or 3) a 2-hour oral glucose tolerance test between 140– 199 mg/dl. T2D controls were defined as adults ≥40 years old who did not meet the definitions of T2D or prediabetes. Some studies, however, used their own in-house definitions and did not identify individuals with prediabetes. Definitions used in the additional biobanks and cohorts were similar to those used in PAGE are summarized in the **Supplemental Information**.

### Genotyping, imputation, and quality control

Details on PAGE genotyping have previously been described.^72^ Briefly, this study included 36,724 PAGE participants who met the T2D and prediabetes definitions and underwent genotyping using the Multi-Ethnic Genotyping Array (MEGA) panel, specifically designed to capture genetic heterogeneity across diverse populations, with the remaining 46,220 PAGE participants genotyped using a variety of other arrays (**Table S1**). Data were imputed using the NHLBI Trans-Omics for Precision Medicine (TOPMed) Imputation Server (https://imputation.biodatacatalyst.nhlbi.nih.gov/#!) using TOPMed-r2 as reference panel, Eagle v2.4 for phasing, and Minimac4 for imputation.^67,73,74^ Variants were excluded if the imputation info score was <0.4 or if the effective sample size was less than 30. The effective sample size was calculated as 2 × MAF × (1-MAF) × N × INFO, where MAF represents the minor allele frequency, N is the sample size, and INFO is the imputation quality score. Principal component analysis was performed separately for each study. Principal components (PCs) were generated using EIGENSOFT^75,76^ as previously described^77^. The top 10 PCs were used to account for population structure in subsequent analyses.

### Construction of T2D PRS

Participants included in this investigation were not part of the discovery GWAS used to develop the T2D PRS. PRS for each participant were constructed as the weighted sum of effect alleles, weighted by variant-specific log ORs using “--score” option with “cols=+scoresums” modifier in PLINK 2.0^78^. All T2D PRS were standardized by subtracting the mean PRS and dividing by the standard deviation of PRS calculated in controls within each population. Therefore, 1 unit increase in PRS corresponds to a 1-standard deviation increase within each population.

The R package “rtracklayer”^79^ was used to align and convert between the GRCh37 (hg19) and the GRCh38 (hg38) genome reference builds across genetic datasets. In the function “GRangesFromDataFrame”, we included a strand field (strand.field= “strand”), which generated the output build with a strand column. If the forward (plus) strand became the reverse (minus) strand in the new build^80^, the strand column would change from “+” to “-”. The alleles were on the new build then changed to the complement of the original input build.

PRS-CSx estimates population-specific and multi-ancestry PRS by jointly modeling discovery GWAS summary statistics and pre-calculated linkage disequilibrium (LD) reference panels based on the 1000 Genomes Project (1KG) Phase 3 participants from AFR, Admixed American (AMR), EAS, EUR, and SAS population samples.^12^ The first three PRS methods used the PRS-CSx “auto” option, which automatically learns the global shrinkage parameter </ from the discovery summary statistics. The META=T option in PRS-CSx was used to generate multi-ancestry PRS using an inverse-variance-weighted meta-analysis of the population-specific posterior effect size estimates (**PRS Method 1 and PRS Method 2**). The META=F option was used to generate five population-specific PRS, which were subsequently calculated in PAGE, standardized to a mean of 0 and SD of 1 by population and study, and used to calculate a linear combination PRS (**PRS Method 3**). In **PRS Method 3**, for each population and study in PAGE, the dataset underwent a three-fold cross-validation process, where the data was evenly divided into three equally sized subsets. In each iteration, one dataset was selected as the testing dataset, while the remaining two were used as the validation dataset. This process was repeated three times to evaluate all possible combinations of validation and testing datasets, so that each individual participated in both the validation and testing sets.

The following logistic regression model of the standardized population-specific PRS was fit in the validation dataset for each population:

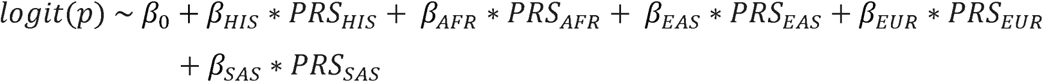

where logit(p) is the log odds of having T2D and *β*_0_ is the intercept, with each subsequent *β* representing the regression coefficient for each population-specific *PRS*. If any of the regression coefficients were <0, the corresponding population-specific PRS and regression coefficient were removed from the regression model, and the model was refitted until all remaining coefficients were >0. The regression coefficients learned in the validation dataset were then used as weights (*w*_*k*_) in the testing datasets to calculate the final linear combination PRS for each individual:

### Final linear combination PRS

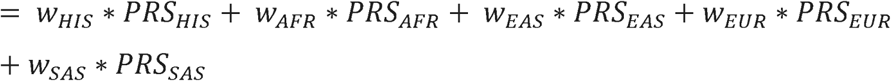

We assessed the predictive accuracy of the four T2D PRS within each population in PAGE using AUCs. AUC values were calculated using the R package “pROC”^81^ with logistic regression of T2D risk after adding PRS to a base model of age, sex, BMI, the first ten PCs of ancestry, and study. The best-performing PRS was calculated in the additional biobanks and cohorts (that were independent of the discovery GWAS used to develop the best-performing PRS) and was used for all subsequent analyses.

### Stratification analyses

We investigated the context-dependent effects of the best-performing T2D PRS in analyses stratified by demographic characteristics and medical history (age group, sex, family history (FH) of T2D, and blood pressure categories), behavioral and lifestyle factors (obesity status, current drinking status, smoking status, and physical activity level), medication use (statin, NSAID, fibrates, lipid-lowering medication, and anti-hypertensive medication), and lipid traits (HDL, LDL, triglycerides, total cholesterol (TC), and non-HDL quartiles) (definitions provided in **Table S5)**. Analyses were conducted in all populations combined and separately in each population using logistic regression models that predict T2D status adjusting for age, sex (except in sex-stratified analyses), BMI, the first ten PCs, and study. AUCs were also calculated using similar models within each stratum. The Cochrane Q-test was used to determine whether effect heterogeneity between subgroups was sufficiently large enough to be attributed to factors beyond sampling error. The P-value from the Cochrane Q-test is referred to as P_het_. A P<0.05 threshold was used to determine statistical significance for stratification analyses.

### T2D PRS associations with diabetes-related traits

We next examined the ability of the best performing T2D PRS to predict 20 traits and complications, including glycemic traits (fasting glucose, fasting insulin, HOMA-IR, and HbA1c), cardiometabolic traits (HDL, LDL, triglycerides, TC, non-HDL, systolic and diastolic blood pressure, and hypertension), vascular diseases (heart attack and stroke), kidney diseases and functions (end stage renal disease, chronic kidney disease, urine albumin-creatine ratio, and eGFR), and inflammatory biomarkers (CRP and IL6) (**Table S10)**. Analyses were performed in all populations combined and separately in each population using regression models adjusting for age, sex, BMI, the top ten PCs, and study. Analyses of glycemic traits were restricted to T2D controls to mitigate confounding effects from diabetes medications in T2D cases. For the rest of the traits, analyses were conducted separately in T2D cases, T2D controls, and prediabetes individuals. Model performance was evaluated using AUCs and odds ratios (ORs) for dichotomous traits and the proportion of variance explained (R^2^) and beta coefficients for continuous traits. Statistical significance was defined as *P* < 2.50 × 10^-3^ (0.05/20 traits).

### PheWAS

To further understand the clinical implication of T2D PRS on the risk of other diseases and conditions, we conducted phenome-wide association studies (PheWASs) in five independent biobanks: BioMe, BioVU, CCPM, MVP, and MGI. Utilizing the PheWAS R package^82^, we ran logistic regression models to assess PRS-phenotype associations, adjusting for age, sex, BMI, and the first ten PCs. For the mapping of the International Classification of Diseases (ICD) 9 and ICD-10 codes to phecodes, we utilized phecodes version 1.2 for ICD-9-CM and the 2018 beta version of ICD-10-CM. Cases were considered valid if they had a minimum phecode count of two. PheWAS analyses were performed separately in each population and study, and a pooled-sample PheWAS was conducted with adjustments for populations in each study. Only phecodes with at least 10 cases within each stratum were considered. A total of 1,815 unique phenotypes across 17 disease categories were tested across the five biobanks, with 1,777, 1,171, 1,813, and 1,695 phenotypes available in AFR, ASN, EUR, and HIS populations, respectively. Significance was defined by correcting for the total number of phenotypes tested in each population using the Bonferroni-adjusted P-value thresholds: *P* < 2.75 × 10^-5^ for all populations combined, *P* < 2.81 × 10^-5^ for AFR, *P* < 4.27 × 10^-5^ for ASN, *P* < 2.76 × 10^-5^ for EUR, and *P* < 2.95 × 10^-5^ for HIS populations. Any *P*-values smaller than the smallest non-zero value that R can represent are denoted as <5.0E-324.

### Meta-analysis

Results from PAGE and the 13 additional biobanks and cohorts were first meta-analyzed using inverse variance weighted fixed-effect models within populations to obtain population-specific results for AFR, ASN, EUR, and HIS populations. The results from these four populations along with the results from all other populations (AI, Greenlandic, MENAQ, and NH), were included in the larger meta-analysis conducted among all populations combined. PheWAS results were similarly meta-analyzed across the five biobanks for each of the four populations and across all populations combined. The degree of heterogeneity was evaluated using the τ2 (between-study variance) and 1^2^ (total proportion of variance owing to heterogeneity) statistics. Meta-analyses were conducted only for analyses with at least 100 cases and 100 controls. If only one study was available for an analysis, making it unfeasible to meta-analyze with other studies, we presented results from that single study.

All statistical analyses were performed using R (version 4.1.2).

## Data availability

The Ge et al. T2D PRS and the T2D PRS of 582 variants are available on the PGS Catalog (https://www.pgscatalog.org; PGS IDs: PGS002308 and PGS000804, respectively). The newly generated PRS in this study will be published on the PGS Catalog. GWAS summary statistics from the Mahajan et al. 2022 study are available on the DIAGRAM consortium website (http://diagram-consortium.org). GWAS summary statistics from the Vujkovic et al., 2020 study are available on dbGaP (accession numbers pha004945.1, pha004943.1, pha004946.1, and pha004944.1, respectively). LD matrices used for PRS-CSx are available at https://github.com/getian107/PRScsx.

## Contributions

B.F.D., H.M.H., S.R., and R.A.J.S. contributed to study conception and design. B.G. and B.F.D. wrote the manuscript. C.K., C.G., and K.E.N. contributed to funding acquisition. Central data analysis group: B.G. (PAGE), D.K. (CCHC), Z.W. (BioMe), J.H. (AoU), R.T. (BioVU), K.A.B. (CLHNS), Y.W. (CHNS), N.P. (CCPM), M.L. (CCPM), F.F.S. (Greenlandic), C.Y. (MESA), B.V. (MGI), K.R.I. and A.T.H. (MVP), N.A.Y. (Qatar), and A.D.P. (REGARDS) contributed to data analysis and interpretation. Study level principal investigators: K.L.M. (CLHNS), P.G.-L. (CHNS), A.A. (Greenlandic), M.B. (MGI), S.S.R. (MESA), A.M. (MESA), J.I.R. (MESA), R.M.I (REGARDS), C.G. (CCPM), K.E.N. (CCHC)., R.J.F.L. (BioMe), T.L.A. (MVP), C.A.H., L.L.M., and L.R.W. (MEC), and C.K. (WHI) contributed data and provided resource and administrative support. B.F.D. and H.M.H. supervised the work. B.F.D. takes responsibility for the integrity and the accuracy of the manuscript. All authors critically reviewed and approved the final version of the manuscript.

## Supporting information

Acknowledgements

