## Supplementary Figures for "Type 2 diabetes polygenic risk score demonstrates context-dependent effects and associations with type 2 diabetes-related risk factors and complications across diverse populations"

**Figure S1**. Density plots of the distribution of T2D PRS by self-reported race and ethnicity populations in the PAGE Study.

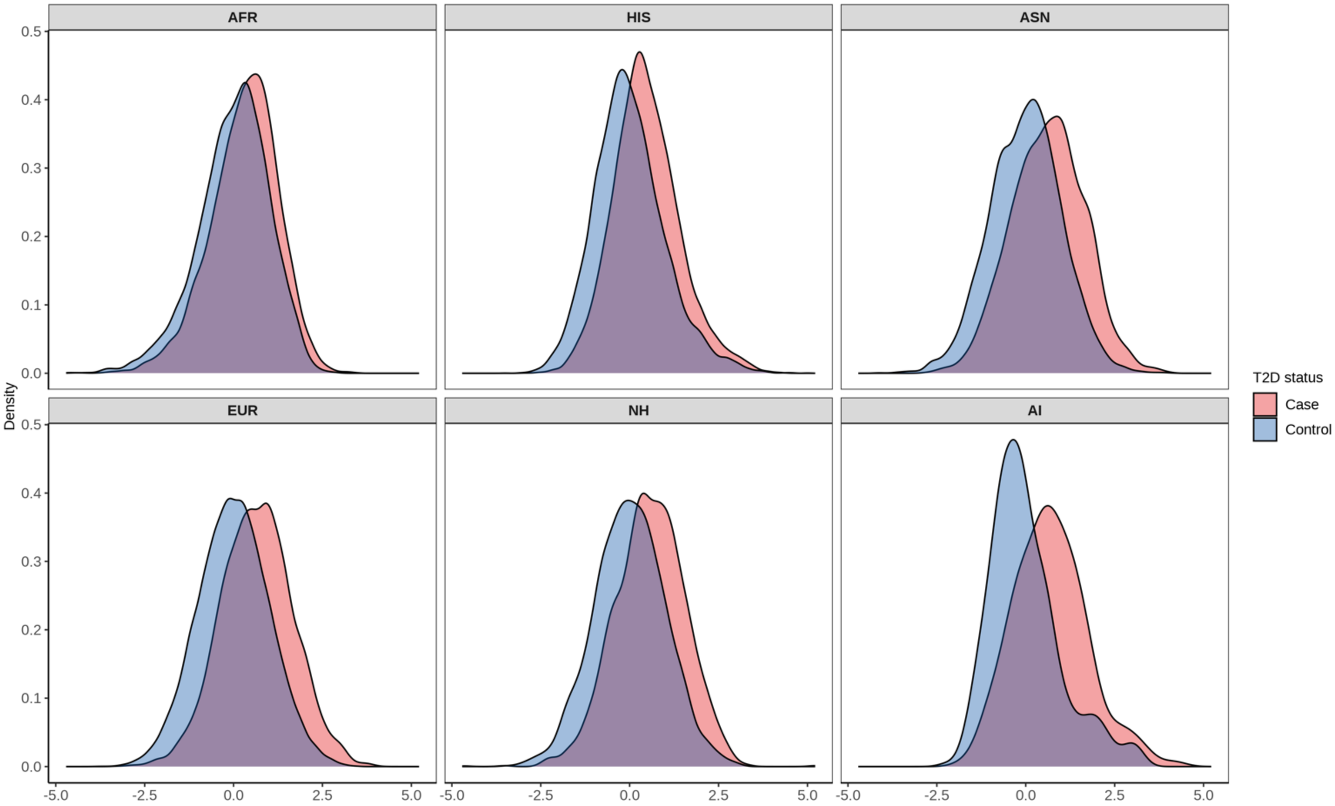


**Figure S2**. Association between each T2D PRS deciles and T2D risk by self-reported race and ethnicity populations in the PAGE Study, using the 40%-60% PRS category as the reference.

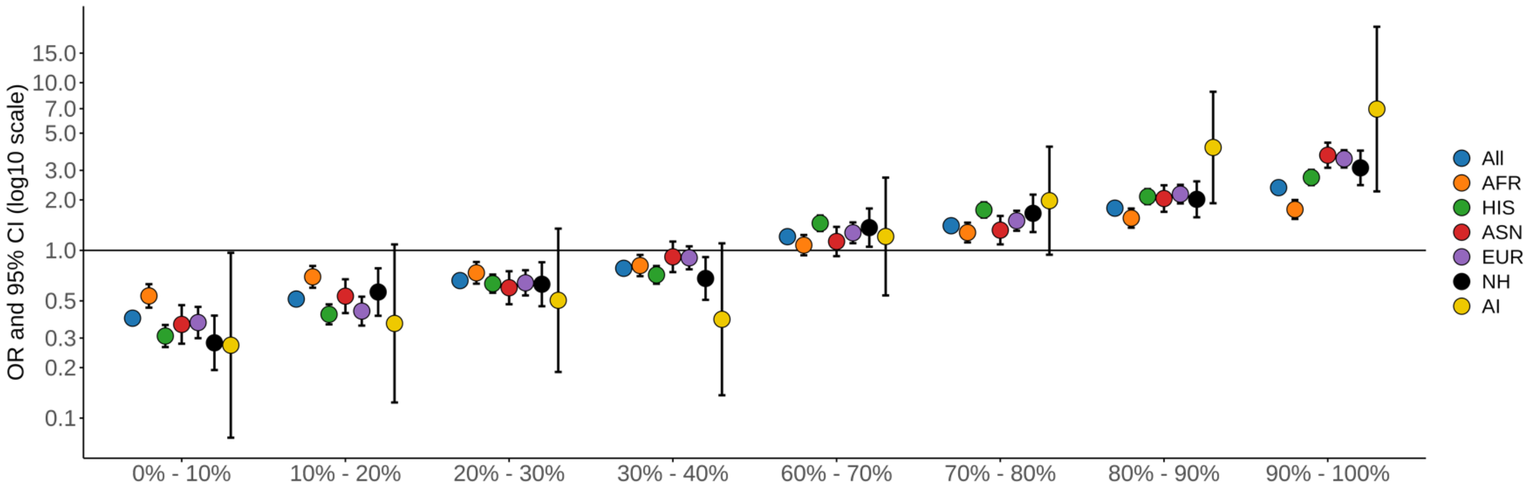

**Figure S3**. Effect of the T2D PRS on T2D risk stratified by demographic, medical, and lifestyle and behavioral factors in the PAGE Study and meta-analyzed results from the additional biobanks and cohorts. P-values are indicated when differences were statistically significant. A) Demographic characteristics and medical history factors. B) Behavioral and lifestyle factors. C) Medication use. D) Lipids.

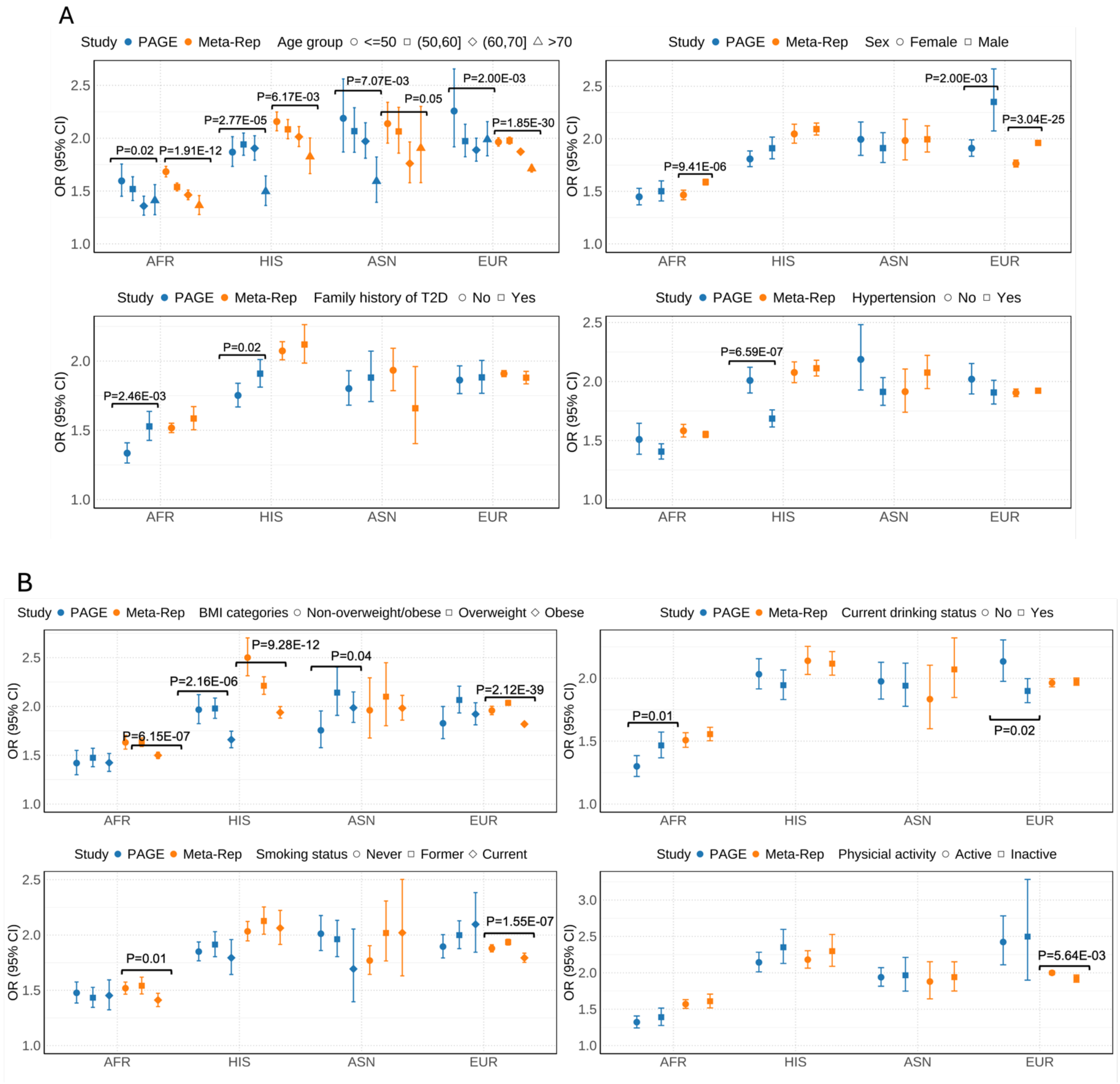

C

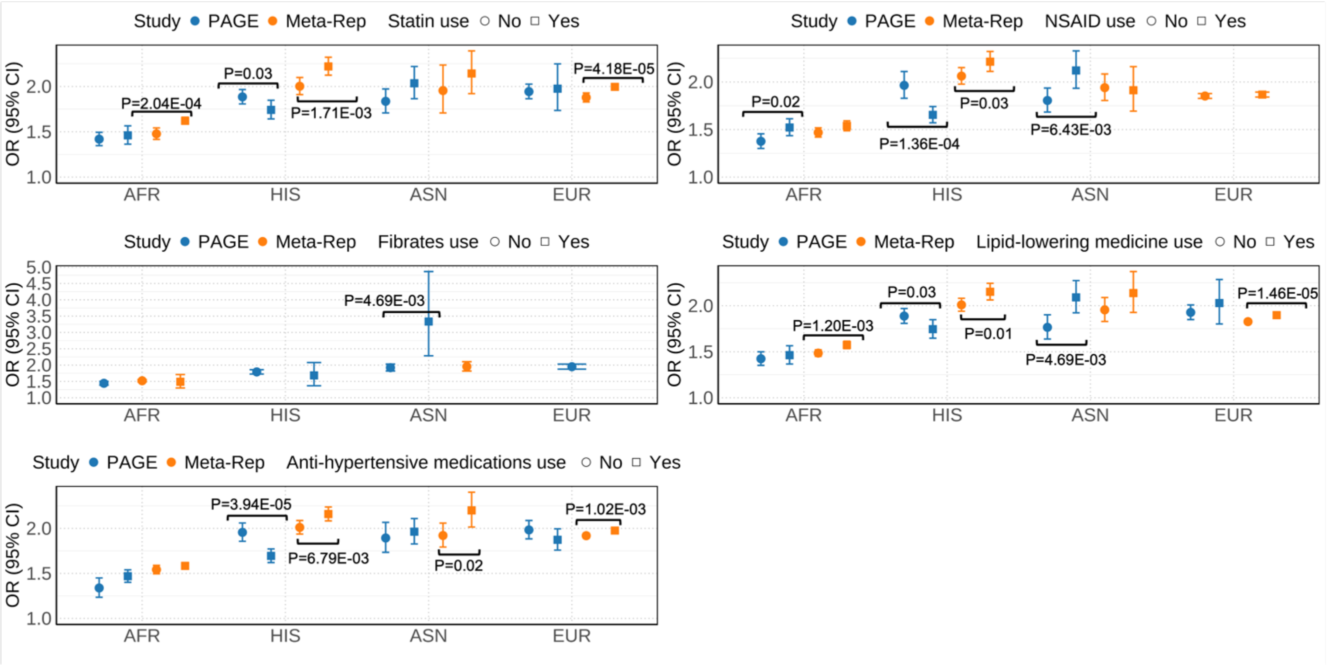
Study PAGE Meta-Rep Statin use No Yes
P=2.04E-04
P=0.03
P=1.71E-03
P=4.18E-05
Study PAGE Meta-Rep NSAID use No Yes
P=0.02
P=1.36E-04
P=0.03
P=6.43E-03
Study PAGE Meta-Rep Fibrates use No Yes
P=4.69E-03
Study PAGE Meta-Rep Lipid-lowering medicine use No Yes
P=1.20E-03
P=0.03
P=0.01
P=4.69E-03
P=1.46E-05
Study PAGE Meta-Rep Anti-hypertensive medications use No Yes
P=3.94E-05
P=6.79E-03
P=0.02
P=1.02E-03
OR (95% CI)
AFR
HIS
ASN
EUR

D

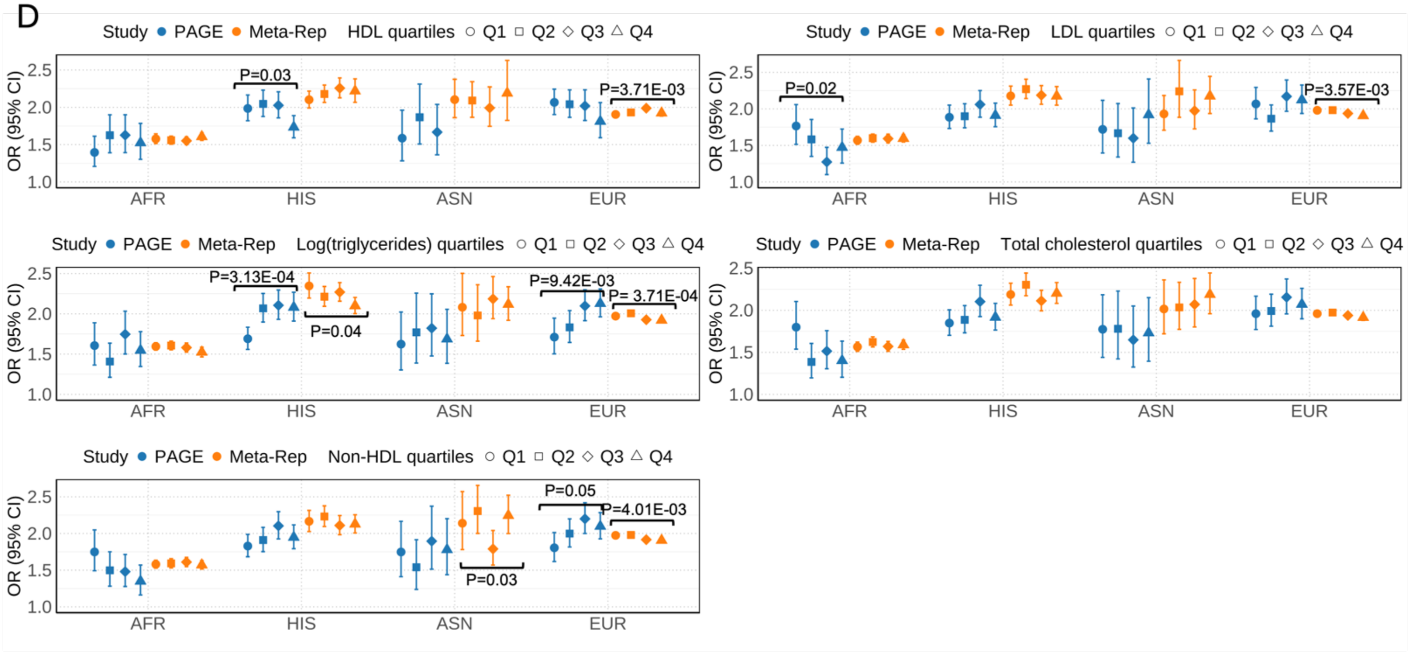
Study PAGE Meta-Rep HDL quartiles Q1 Q2 Q3 Q4
P=0.03
P=3.71E-03
Study PAGE Meta-Rep LDL quartiles Q1 Q2 Q3 Q4
P=0.02
P=3.57E-03
Study PAGE Meta-Rep Log(triglycerides) quartiles Q1 Q2 Q3 Q4
P=3.13E-04
P=0.04
P=9.42E-03
P= 3.71E-04
Study PAGE Meta-Rep Total cholesterol quartiles Q1 Q2 Q3 Q4
Study PAGE Meta-Rep Non-HDL quartiles Q1 Q2 Q3 Q4
P=0.03
P=0.05
P=4.01E-03
OR (95% CI)
AFR
HIS
ASN
EUR

**Figure S4.** T2D PRS PheWAS results meta-analyzed across all five biobanks by population. The X-axis represents phecodes color-coded by their corresponding phenotype category and is ordered from the category with the most to the category with the least significant hits. The Y-axis represents the $-\log_{10}$(p-value). The red horizontal line represents Bonferroni-adjusted p-value thresholds (EUR: $P < 2.76 \times 10^{-5}$, HIS: $P < 2.95 \times 10^{-5}$ AFR: $P < 2.81 \times 10^{-5}$and ASN: $P < 4.27 \times 10^{-5}$ ), and the blue horizontal line represents an unadjusted p-value threshold of $P < 0.05$. Upward triangles indicate positive associations, while downward triangles indicate negative associations. The top ten most significant associations from the endocrine/metabolic category are annotated, while the single most significant association from each other category is annotated.

A) EUR population

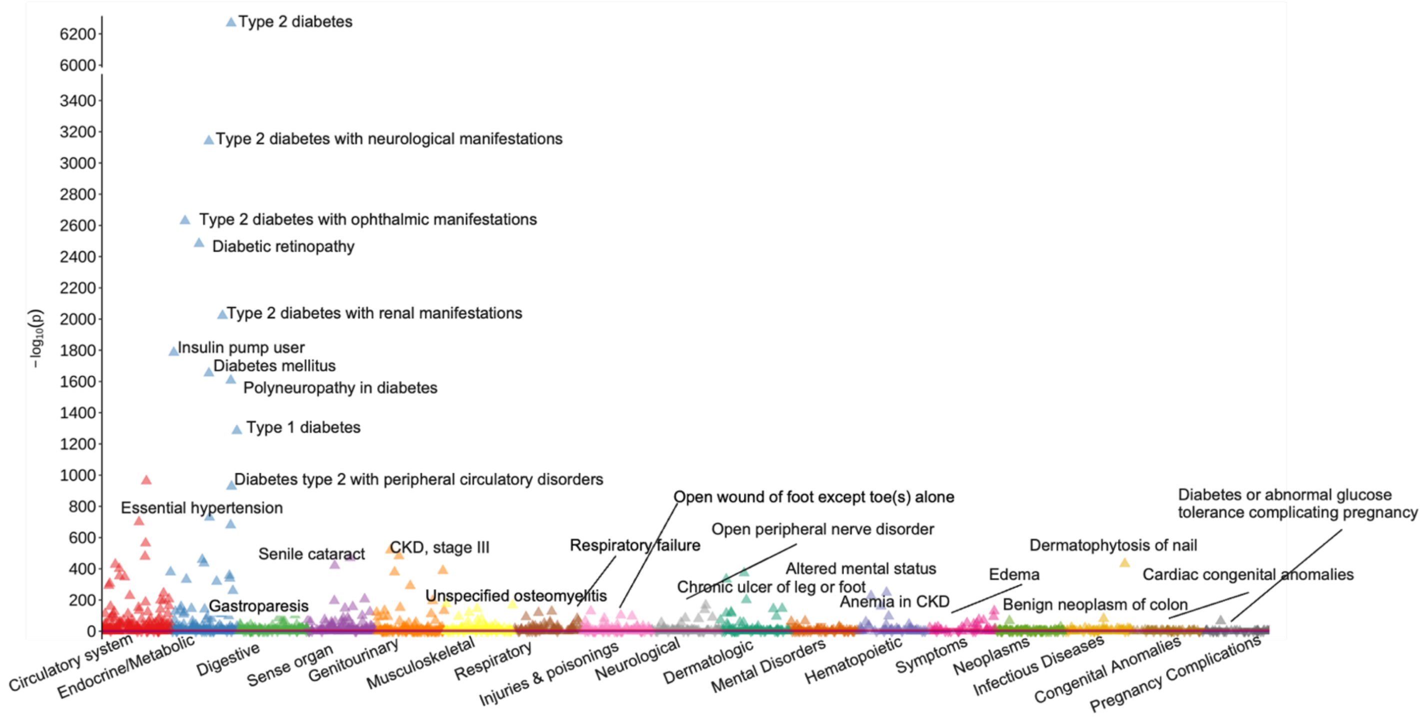

B) HIS population

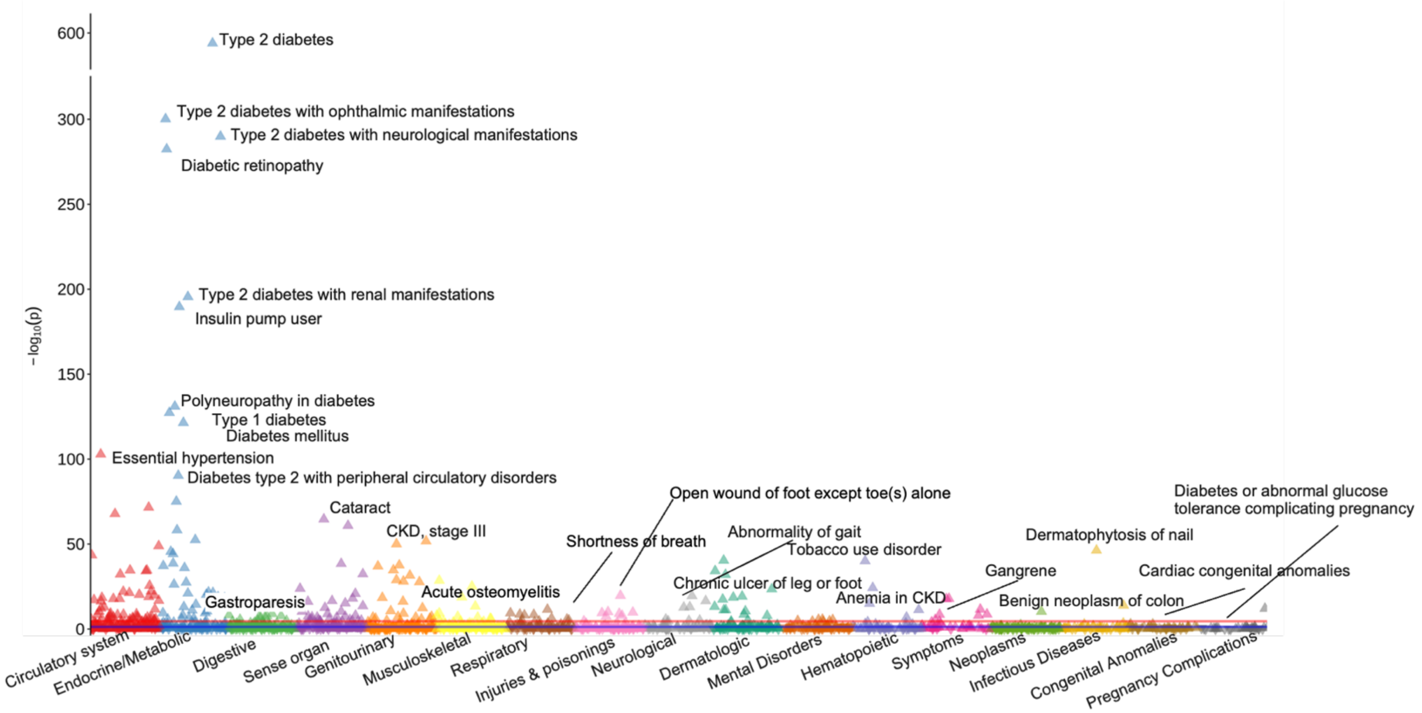


C) AFR population

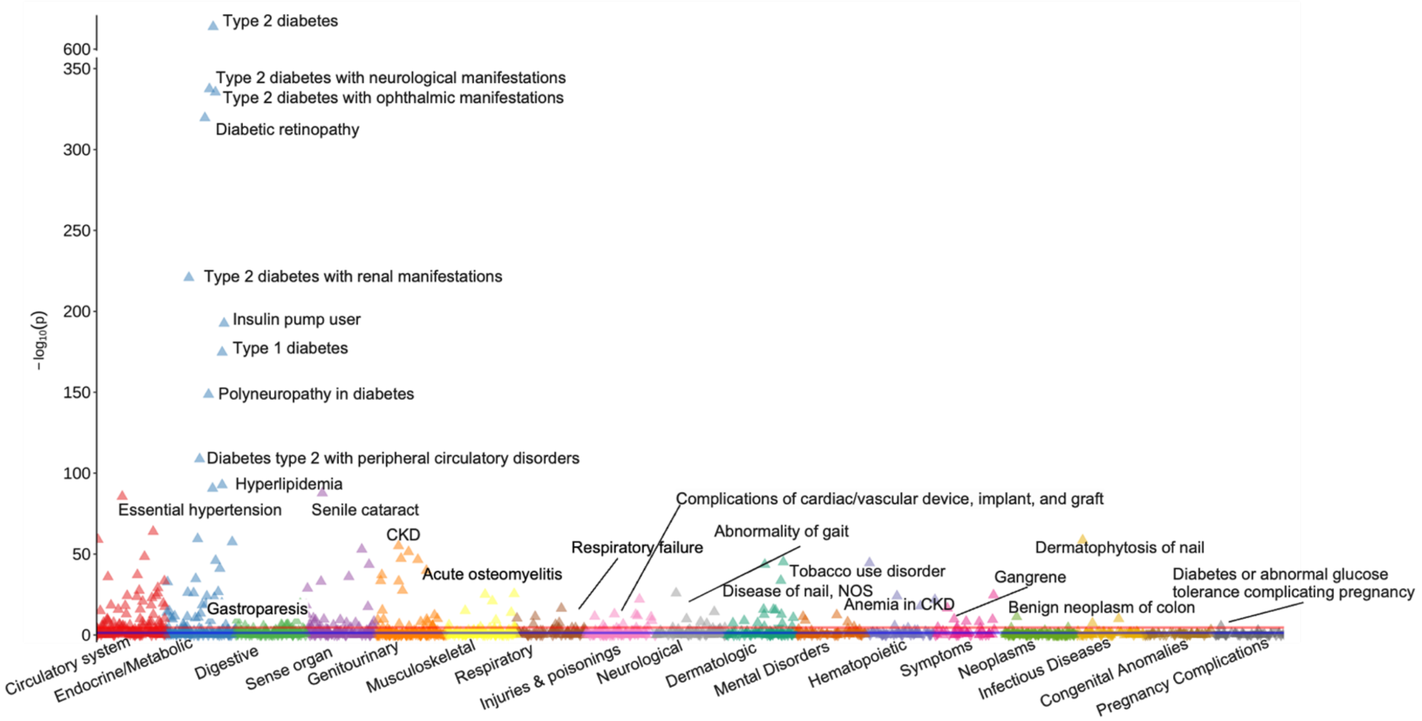

D) ASN population

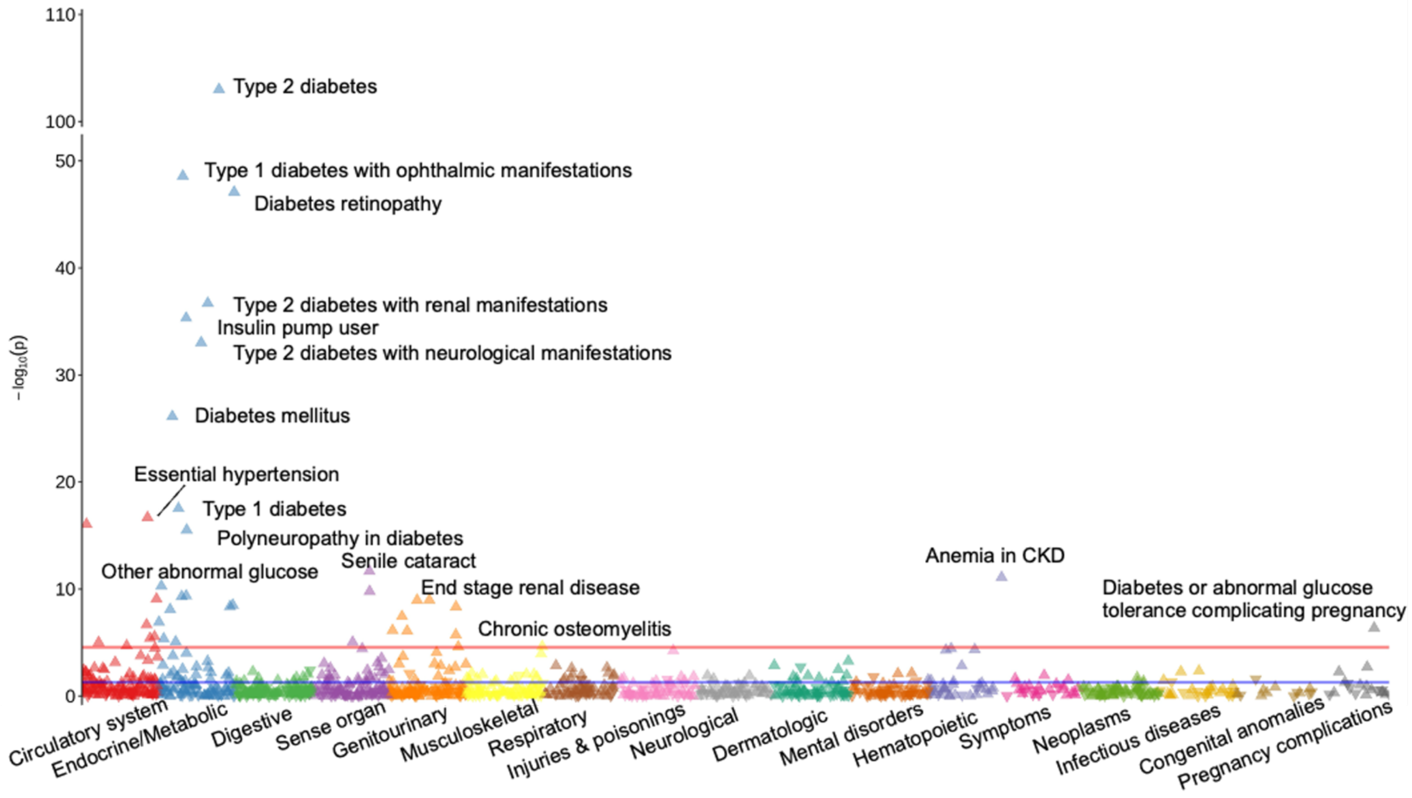
