## Supplementary Information for "Type 2 diabetes polygenic risk score demonstrates context-dependent effects and associations with type 2 diabetes-related risk factors and complications across diverse populations"

**Description of the included studies, and definitions of T2D cases, individuals with prediabetes, and T2D controls**

I. **PAGE-participating studies**

**Atherosclerosis Risk in Communities (ARIC)**^1^ is a longitudinal cohort study designed to investigate the etiology of atherosclerosis and its clinical outcomes. Initiated in 1987, the ARIC study recruited participants between the ages of 45 and 65 from four U.S. communities: Forsyth County, NC; Jackson, MS; Washington County, MD; and Minneapolis, MI. Participants received an extensive examination of their medical, social and demographic profiles approximately every three years.

In the ARIC study, blood samples were collected during the baseline visit. Prevalent T2D was identified during the baseline examination if participants exhibited ≥8 h fasting blood glucose levels ≥126 mg/dL, or non-fasting glucose ≥200 mg/dL, self-reported a physician diagnosis of diabetes or “sugar in the blood,” or reported current use of diabetes medication within the last two weeks. Participants were defined as T2D controls if they did not report using diabetes medication within the past two weeks and had fasting glucose levels <126 mg/dL.

**The Icahn School of Medicine at Mount Sinai BioMe biobank in New York City (BioMe)** is an electronic medical record (EMR)-linked biobank enriched for racially and ethnically diverse populations recruited from Mount Sinai Medical Center in upper Manhattan area, New York City since 2007. Data on anthropometrics, demographics, and medication use were derived from participants’ EMR and a medical history questionnaire administered at baseline. (<https://icahn.mssm.edu/research/ipm/programs/biome-biobank/facts>).

BioMe implemented the eMERGE algorithm^2^ to define T2D cases and controls. T2D cases were participants meeting any of the following criteria: 1) having International Classifiers of Disease (ICD-9-CM) codes of 250.x0 or 250.x2, except for 250.10 and 250.12, 2) on T2D medications and/or insulin, or 3) glucose > 200 mg/dl or HbA1c ≥ 6.5% laboratory results. Cases with a T2D diagnosis code were required to have either an abnormal laboratory test or a T2D medication prescription. Cases without a T2D diagnosis code were required to have documentation of both a prescription for a T2D medication and random glucose >200 mg/dl, or fasting glucose >125 mg/dl or HbA1c ≥6.5%. For patients treated with insulin alone, past prescription for a T2D medication or meeting specific criteria (no T1D diagnoses and ≥ 2 T2D diagnosis dates) were necessary.

T2D controls were defined by excluding those with any diabetes diagnosis (ICD-9-CM codes 250.xx), patients on insulin or any of diabetes medications, patients who used any diabetic supplies, and patients with any abnormal glucose (≥110 mg/dl) or HbA1c (≥6.0%) values. Patients with a family history of diabetes were also excluded. Controls were required to have ≥ 1 normal glucose measurement and ≥ 2 in-person clinician encounters.

**Coronary Artery Risk Development in Young Adults (CARDIA)**^3^ is a longitudinal cohort study aims to identify factors that contribute to cardiovascular diseases in later life, and to better understand the natural history of cardiovascular diseases over the adult life. 5,115 African American and European American participants aged 18 to 30 years were recruited from four urban communities: Birmingham, AL; Chicago, IL; Minneapolis, MN; and Oakland, CA in 1985-1986. In-person examinations occurred at 2, 5, 7, 10, 15, 20, 25, and 30 (2015-2016) years after baseline with annual telephone interviews to update health status and contact information.

T2D cases were identified at each examination using a combination of available information on ≥126 mg/dL fasting glucose, postchallenge (75 g) glucose ≥200 mg/dL, HbA1c ≥6.5%, or use of diabetes medications. Individuals diagnosed at <20 years old were excluded as they were considered to have T1D. T2D controls were defined as fasting glucose levels (<7.0 mmol/L and <126 mg/dL) at last available visit or no report of diabetes medication^4^.

**Hispanic Community Health Study/Study of Latinos (HCHS/SOL)**^5,6^ is a longitudinal cohort study of adults who were self-identified Hispanic/Latino and aged 18 to 74 years living in four study sites – Bronx, NY; Chicago, IL; Miami, FL; and San Diego, CA. The HCHS/SOL aims to determine the role of acculturation in the prevalence and incidence of diseases, and to identify influencing factors for the health of Hispanic/Latino populations. Participants received an extensive clinic exam and assessments to determine baseline risk factors during 2008-2022, had a second clinical visit during 2014-2017, and the third visit in January 2020 and concluded in January 2024.

T2D cases were defined as individuals with fasting time >8 h and fasting glucose levels ≥126 mg/dL, fasting ≤8 h and fasting glucose ≥200 mg/dL, post–oral glucose tolerance test glucose ≥200 mg/dL, HbA1c ≥6.5%, or on current treatment with antihyperglycemia medications. T2D controls were those with fasting time >8 h and fasting glucose levels <100 mg/dL, post–oral glucose tolerance test glucose <140 mg/dL, and HbA1c <5.6%^7^.

**Multiethnic Cohort Study (MEC)**^8^ is a longitudinal cohort study which follows over 215,000 residents of Hawaiʻi and Los Angeles for development of cancer and other chronic diseases. It includes men and women of five main racial and ethnic groups, Japanese Americans, Native Hawaiians, African Americans, Latinos, and whites. The participants aged 45 to 75 years were recruited between 1993 and 1996. Biological specimens were collected from more than 70,000 MEC members from 2001 to 2005. Eight sub-studies from MEC were also included in this manuscript. They are MEC-AABC (African Ancestry Breast Cancer), MEC-AAPC (African Ancestry Prostate Cancer), MEC-JABC (Japanese Ancestry Breast Cancer), MEC-JAPC (Japanese Ancestry Prostate Cancer), MEC-LABC (Breast Cancer in Latinos), MEC-LAPC (Prostate Cancer in Latinos), MEC-SIGMA (the Slim Initiative in Genomic Medicine for the Americas), and MEC-HIBC (Native Hawaiian Breast Cancer).

T2D cases were defined as: 1) a self-report of diabetes on the baseline questionnaire, 2nd questionnaire or 3rd questionnaire, or 2) self-report of T2D medication at the time of blood draw, or 3) no diagnosis of T1D in the absence of a T2D diagnosis from the California Office of Statewide Health Planning and Development (OSHPD) for California Residents, or 4) individuals who were linked to the diabetes registries of Hawaii Medical Service Association (HMSA) or Kaiser Permanente (KPH) Hawaii health plans, or who were designated as diabetic in the Chronic Conditions Data Warehouse of Medicare ^9^. T2D controls were defined as: 1) no self-report of diabetes on any of the questionnaires while having completed a minimum of 2 of the 3 (79% of controls returned all 3 questionnaires); and 2) no use of medications for T2D at the time of blood draw; and 3) no T1D and T2D diagnosis from the OSHPD, HMSA or KPH registries. To preserve DNA for genetic studies of cancer in the MEC, subjects with an incident cancer diagnosis at time of selection for this study were excluded. Controls were frequency matched to cases on age at entry into the cohort (5-year age groups) and for Latinos, place of birth (U.S. vs. Mexico, South or Central America), oversampling African American, Native Hawaiian and European American controls to increase statistical power. For controls with glucose measured, individuals with >126 mg/dL were also excluded.

**Women’s Health Initiative (WHI)**^10^ is a longitudinal cohort study focuses on strategies for preventing heart disease, breast and colorectal cancer, and osteoporosis in postmenopausal women. The WHI study collected long-term data from 52,068 WHI women aged 50 – 79 years between 1993 and 1998 at 40 clinical centers across the U.S. Seven sub-studies from WHI were also included in this manuscript. They are WHI-GARNET(Genomics and Randomized Trials Network), WHI-HIPFX (the Hip Fracture GWAS), WHI-GECCO (the Genetics and Epidemiology of Colorectal Cancer Consortium), WHI-LLS (the Long Life Study), WHI-MOPMAP (the Modification of PM-Mediate Arrhythmogenesis in Population study), WHI-WHIMS (Women’s Health Initiative Memory Study), and WHI-SHARe (SNP Health Association Resource).

T2D was documented at baseline by self-report in which each woman was asked whether she had ever been told that she had “sugar diabetes” by her physician. Incident T2D cases were identified during annual follow-up with self-administered questionnaires. A WHI diabetes confirmation study has demonstrated consistency between these medical inventories and incident and prevalent diabetes^11^. T2D controls were selected if participants reported no diabetes at baseline and did not self-report of medication to treat diabetes through their last follow-up.

II. **Additional biobanks and cohorts**

**All of Us (AoU)**^12^ is a longitudinal cohort study aims to collect data from at least one million individuals across the United States, creating a diverse health database for epidemiological and genomic studies. Initiated in May 2018, AoU has recruited over 781,000 participants as of March 26, 2024.

We defined T2D cases in AoU using the following criteria: adults aged ≥ 25 years and 1) reported T2D diagnosis, or current T2D treatment and medication use, or 2) had T2D, disorder due to T2D, or had complications due to T2D at ≥2 occurrence dates; or 3) had fasting glucose ≥126 mg/dL, or random glucose ≥200 mg/dL, or HbA1c ≥6.5%, at ≥2 occurrence dates. We excluded 1) those did not have short read whole genome sequencing (WGS) data, 2) indicated T1D diagnosis in the survey, 3) had T1D or related complications and disorder at ≥2 occurrence dates, 4) had pregnancy related T1D/T2D at ≥2 occurrence dates. Individuals with prediabetes were defined as WGS participants who: 1) reported prediabetes diagnosis, treatment and medication use in the survey, or 2) had T1D at ≥2 occurrence dates, or 3) had 100-125 mg/dL fasting glucose, 5.7–6.4% HbA1c, or 140–199 mg/dL from glucose tolerance 2 hours panel at ≥2 occurrence dates. T2D controls were defined as WGS participants aged ≥40 years and did not meet the definitions of T2D cases and prediabetes.

**BioVU Biobank (BioVU)**^13^ is a de-identified EHR-based biobank launched in 2007 at Vanderbilt University Medical Center (VUMC), a tertiary-care center at Nashville, TN. BioVU consents patients in the out-patient clinic environments at VUMC to have their leftover blood from clinical care be banked for future research. BioVU aims to provide a resource for studies of genotype-phenotype associations (https://victr.vumc.org/what-is-biovu/). Owing to the de-identification process all BioVU participants are deemed non-human subjects by the Institutional Review Board.

Three sets of criteria were used to define T2D cases. Case definition I: 1) any of the following ICD-9-CM codes—250.3, 250.32, 250.2, 250.22, 250.9, 250.92, 250.8, 250.82, 250.7, 250.72, 250.6, 250.62, 250.5, 250.52, 250.4, 250.42, 250, and 250.02—or equivalent ICD-10-CM codes and 2) use of non-insulin diabetes medication. Case definition II: 1) any of the above ICD-9 codes; 2) a glucose concentration greater than 200 mg/dL or a HbA1c level greater than 6.5%; and 3) no use of insulin medication. Case definition III: 1) use of non-insulin diabetes medication and 2) a glucose concentration greater than 200 mg/dL or a HbA1c level greater than 6.5%. The controls were defined as “record does not contain any of the following information”: 1) ICD-9/10-CM codes from the case definition or any of following ICD-9-CM codes—790.21, 790.22, 790.29, 791.5, 648.8, 277.7, and 250, or equivalent ICD-10-CM; 2) use of insulin medication; 3) use of non-insulin diabetes medication; 4) history of diabetes; and 5) a glucose concentration ≥110 mg/dL or a HbA1c level ≥6.0%^14^.

**Cameron County Hispanic Study (CCHC)**^15^ is a randomly-ascertained, community-based cohort of study that recruited more than 5,000 Mexican American participants in Brownsville (Cameron County), TX since 2004. CCHC participants were followed at five, 10, and 15 years. Demographic, lifestyle, clinical measurements, and biospecimens were collected at each visit. The analyses were restricted to individuals with no closer than 3rd degree genetic kinship with each other (N=3019) who had measured genotype.

T2D cases were defined as participants who had either 1) T2D diagnosis; or 2) T2D medication use; 3) or fasting blood glucose ≥126 mg/dL; or 4) HbA1C ≥ 6.5%. Individuals with prediabetes were defined as participants without a T2D diagnosis, no diabetes medication use, and either had a fasting blood glucose level ≥100 mg/dL and <126 mg/dL or a HbA1C level ≥ 5.7% and <6%. T2D controls were participants who had no T2D diagnosis, no diabetes medication use, had a fasting blood glucose level <100 mg/dL, and a HbA1C level < 5.7%.

**Cebu Longitudinal Health and Nutrition Survey (CLHNS)**^16^ is an ongoing longitudinal study with a focus on infant feeding patterns among Filipino women who gave birth between May 1, 1983, and April 30, 1984 (<https://cebu.cpc.unc.edu/>). The Cebu study expanded its scope to investigate various health, demographic, and nutritional outcomes, including birth weight, infant feeding practices, maternal diet, and child spacing. The baseline survey included 3,327 pregnant women from the Metropolitan Cebu area, with subsequent surveys conducted bimonthly for 24 months. Follow-up surveys tracked the participants into adolescence and young adulthood, aiming to understand the intergenerational effects on health and nutrition. Overnight fasting blood samples for biomarker measurement and DNA extraction were obtained in 2005.

T2D cases were defined as participants who were currently on T2D medication, and/or those with adjusted glucose level ≥7.0 mmol/L. Adjusted glucose was calculated by subtracting 0.97 mmol/l from the measured overnight fasting blood glucose level. Individuals with prediabetes were defined as participants who did not meet the definition of T2D cases and had a glucose adjusted level ≥100 mg/dL. T2D controls were participants who were not on T2D medication, and had overnight fasting blood glucose <7.0 mmol/L.

**China Health and Nutrition Survey (CHNS)**^17^ is a nationwide longitudinal study aimed at investigating a range of economic, sociological, demographic, and health-related inquiries within the Chinese population. Utilizing a stratified probability sampling method with a multistage, random cluster design, counties and cities across nine diverse provinces (Guangxi, Guizhou, Heilongjiang, Henan, Hubei, Hunan, Jiangsu, Liaoning, and Shandong) were selected, stratified by income and urbanization according to State Statistical Bureau definitions. A total of 4,560 households from 228 communities were then randomly chosen from each stratum. CHNS began in 1989 with additional surveys completed in 1997, 2000, 2004, 2006, 2009, 2011, 2015, and 2018.

CHNS used the same definitions of T2D cases, individuals with prediabetes, and T2D controls as PAGE (refer to the Methods - T2D cases and controls definition section), except that CHNS does not have random glucose and OGTT measurements.

**Colorado Center for Personalized Medicine Biobank (CCPM)**^18^ is a EHR linked biobank that was jointly developed by the University of Colorado Anschutz Medical Campus and University of Colorado Health (UCHealth) to serve as a unique, dual-purpose research and clinical resource accelerating personalized medicine. Began in September 2015, CCPM enrolled >200,000 participants who received medical care at UCHealth University of Colorado Hospital in Denver, CO.

T2D cases were determined through the use of phecode 250.2X sourced from the PheWAS catalog (https://phewascatalog.org/), which maps ICD-9 and ICD-10 codes from CCPM participants to specific phecodes. Case identification required the presence of at least two distinct separate encounters. Exclusions were then applied to identify T2D controls.

**Greenlandic Study (Greenlandic)** consists of two cross-sectional Greenlandic cohorts^19^: The Inuit Health in Transition (IHIT)^20^ and the Greenland population study (B99)^21^. The B99 and the IHIT cohorts were collected as part of a general population health survey of the adult Greenlandic population during 1998–2001 and 2005–2010, respectively. Only participants living in Greenland were included. The Greenlandic population is of mainly Inuit genetic ancestry but also has recent gene flow from Europe (on average 25% European ancestry and 75% Inuit ancestry).

The Greenlandic participants underwent an OGTT in which blood samples were drawn after an overnight fast and after 2 h during a 75g OGTT. The Greenlandic studies used the same definitions of T2D cases, individuals with prediabetes, and T2D controls as PAGE (refer to the Methods - T2D cases and controls definition section).

**Multi-Ethnic Study of Atherosclerosis Study (MESA)**^22^ is a prospective cohort study aimed at studying the prevalence, progression, determinants, and prognostic significance of subclinical cardiovascular disease in a sex-balanced, multiethnic, community-dwelling U.S. cohort. MESA was initiated in 1999 and recruited >6,000 participants aged 45-84 seen at clinics in Columbia University, New York; Johns Hopkins University, Baltimore; Northwestern University, Chicago; UCLA, Los Angeles; University of Minnesota, Twin Cities; and Wake Forest University, Winston Salem (https://www.mesa-nhlbi.org/)

MESA used the same definitions of T2D cases, individuals with prediabetes, and T2D controls as PAGE (refer to the Methods - T2D cases and controls definition section), except that MESA does not have HbA1c and OGTT measurements.

**Michigan Genomics Initiative (MGI)**^23^ is a single health-system biobank comprising >91,000 participants with age ranging from 18 to over 90 years, recruited primarily during surgical encounters at Michigan Medicine. MGI recruitment began in 2012 with the goal of combining patient EHR data with corresponding genetic data to gain novel biomedical insights and to accelerate biomedical and precision health research at the University of Michigan.

T2D cases were identified using the PheWAS R package^24^ to map ICD-10 codes to phecodes (250.2X), and excluded those had a T1D phecode (250.1X). T2D controls were all individuals other than T2D cases.

**Million Veteran Program (MVP)**^25–27^ is a cohort of fully consented veterans from the United States military, gathered from more than 75 participating Department of Veterans Affairs (VA) medical facilities. Since recruitment efforts started in 2011, 1 million Veterans have joined MVP with genotype data on over 650,000 racially/ethnically diverse individuals. Each study participant contributed blood samples for DNA extraction and genotyping, in addition to completing surveys regarding their health status, lifestyle choices, and military service experiences. Informed consent is also obtained from all participants to access to their full EHR within the VA prior to and after enrollment including inpatient International Classification of Diseases (ICD9/10) diagnosis codes, Current Procedural Terminology (CPT) codes, clinical laboratory measurements, and reports of diagnostic imaging modalities. The EHR is continuously being integrated with MVP genomic data and access to these linked coded data is provided to approved investigators. The study received ethical and study protocol approval from the VA Central Institutional Review Board.

Prevalent T2D cases were identified using precedes 250.2X, requiring the presence of at least two separate encounters at or before enrollment. T2D controls were all individuals other than T2D cases.

**Qatar Biobank (Qatar)**^28^ is a population-based research initiative that creates a repository of biological samples and information on health and lifestyle of Qatari citizens and long-term residents (<https://www.qatarbiobank.org.qa/>). The Qatar Biobank aims to collect extensive lifestyle, clinical, and biological information from up to 60,000 Qatari nationals and long-term residents (individuals living in the country for ≥15 years) aged ≥18 years (approximately one-fifth of all Qatari citizens). Consented participants underwent a comprehensive assessment at the Qatar Biobank facility in Doha, Qatar. The assessment consisted of a 5-stage interview and physical examination sequence, lasting approximately 3 hours on average. Participants provided biological samples (blood, urine, and saliva), which were analyzed at diagnostic laboratories.

T2D cases were identified if 1) they answered yes to the question "Has a doctor ever told you that you had or have diabetes?", or 2) if their HbA1c level was ≥6.5%, or 3) if fasting glucose level ≥126 mg/dL or non-fasting glucose level ≥200 mg/dL. Those took insulin were excluded as they were considered to have T1D. Additionally, individuals with prediabetes, defined as having fasting glucose levels between 100-125 mg/dL or HbA1c levels between 5.7 and 6.4, were removed, with precedence given to a diagnosis of diabetes over prediabetes. Furthermore, individuals diagnosed with T2D at age <32 years, suspected of having T1D, were also excluded. Any remaining individuals not meeting these criteria were designated as T2Dcontrols.

**REasons for Geographic and Racial Differences in Stroke Study (REGARDS)**^29^ is a national observational study aimed at investigating factors contributing to stroke risk in adults aged 45 years or older. Between January 2003 and October 2007, 30,239 participants were recruited and underwent a telephone interview followed by an in-home physical examination. Assessments included traditional stroke risk factors such as blood pressure and cholesterol levels, as well as electrocardiograms of the heart. Participants are contacted every six months via phone to inquire about stroke symptoms, hospitalizations, and general health status.

T2D cases were defined at the baseline and second in-home visits as fasting glucose ≥126 mg/dL or a random glucose ≥200 mg/dL among those who did not fast, or self-reported use of insulin or oral diabetes medication. T2D controls were everyone else who were not missing relevant data to classify the phenotype.
