## Supplementary material for "Type 2 diabetes polygenic risk score demonstrates context-dependent effects and associations with type 2 diabetes-related risk factors and complications across diverse populations": Acknowledgements

The **Population Architecture Using Genomics and Epidemiology (PAGE)** program is funded by the National Heart, Lung and Blood Institute (NHLBI), supported by R01HL151152.

The contents of this paper are solely the responsibility of the authors and do not necessarily represent the official views of the NIH. The PAGE consortium thanks the staff and participants of the PAGE studies for their important contributions. The listing of PAGE senior investigators can be found at <http://www.pagestudy.org>.

**PAGE-participating studies**

**ARIC**: The Atherosclerosis Risk in Communities study has been funded in whole or in part with Federal funds from the National Heart, Lung, and Blood Institute, National Institutes of Health, Department of Health and Human Services (contract numbers HHSN268201700001I, HHSN268201700002I, HHSN268201700003I, HHSN268201700004I and HHSN268201700005I). The authors thank the staff and participants of the ARIC study for their important contributions. The datasets used for the analyses described in this manuscript were obtained from dbGaP under accession phs000223. We also acknowledge R01 HL143885 for multiomics, metabolomics, obesity and cardiovascular disease research.

**BioMe**: The Mount Sinai BioMe Biobank is supported by The Andrea and Charles Bronfman Philanthropies and by Federal funds from the NIH (U01HG00638001; U01HG007417; X01HL134588). Furthermore, analyses were in part supported through the computational and data resources and staff expertise provided by Scientific Computing and Data at the Icahn School of Medicine at Mount Sinai and by the Clinical and Translational Science Awards (CTSA) grant UL1TR004419 from the National Center for Advancing Translational Sciences. Research reported in this publication was also supported by the Office of Research Infrastructure of the National Institutes of Health under award number S10OD026880 and S10OD030463. The content is solely the responsibility of the authors and does not necessarily represent the official views of the National Institutes of Health.

The data used for the analyses described in this manuscript were obtained from dbGaP under accession phs000925.

**CARDIA**: The Coronary Artery Risk Development in Young Adults Study (CARDIA) is conducted and supported by the National Heart, Lung, and Blood Institute (NHLBI) in collaboration with the University of Alabama at Birmingham (HHSN268201800005I & HHSN268201800007I), Northwestern University (HHSN268201800003I), University of Minnesota (HHSN268201800006I), and Kaiser Foundation Research Institute (HHSN268201800004I). CARDIA was also partially supported by the Intramural Research Program of the National Institute on Aging (NIA) and an intra-agency agreement between NIA and NHLBI (AG0005).. The data used for the analyses described in this manuscript were obtained from dbGaP under accession phs000236.We also acknowledge R01 HL143885 for multiomics, metabolomics, obesity and cardiovascular disease research.

**HCHS/SOL**: The Hispanic Community Health Study/Study of Latinos is a collaborative study supported by contracts from the National Heart, Lung, and Blood Institute (NHLBI) to the University of North Carolina (HHSN268201300001I / N01-HC-65233), University of Miami (HHSN268201300004I / N01-HC-65234), Albert Einstein College of Medicine (HHSN268201300002I / N01-HC-65235), University of Illinois at Chicago – HHSN268201300003I / N01-HC-65236 Northwestern Univ), and San Diego State University (HHSN268201300005I / N01-HC-65237). The following Institutes/Centers/Offices have contributed to the HCHS/SOL through a transfer of funds to the NHLBI: National Institute on Minority Health and Health Disparities, National Institute on Deafness and Other Communication Disorders, National Institute of Dental and Craniofacial Research, National Institute of Diabetes and Digestive and Kidney Diseases, National Institute of Neurological Disorders and Stroke, NIH Institution-Office of Dietary Supplements. The data used for the analyses described in this manuscript were obtained from dbGaP under accession phs000555.

**MEC**: The MEC study is funded through the National Cancer Institute (U01 CA164973). The datasets used for the analyses described in this manuscript were obtained from dbGaP under accession phs000220.

**WHI**: The WHI program is funded by the National Heart, Lung, and Blood Institute, National Institutes of Health, U.S. Department of Health and Human Services through contracts 75N92021D00001, 75N92021D00002, 75N92021D00003, 75N92021D00004, 75N92021D00005. The datasets used for the analyses described in this manuscript were obtained from dbGaP under accession phs000227.

**Additional biobanks and cohorts**

**BioVU**: Vanderbilt University Medical Center’s BioVU projects are supported by numerous sources: institutional funding, private agencies, and federal grants. These include National Institute of Health funded Shared Instrumentation Grant S10OD017985, S10RR025141, and S10OD025092; Clinical and Translational Science Awards grants UL1TR002243, UL1TR000445, and UL1RR024975. Data curation and phenotyping in this project are supported by SFORN grant 17SFRN33520017 from the American Heart Association.

Ethics approval and consent to participate

BioVU Consent form is provided to patients in the outpatient clinic environments at VUMC. The consent states policies on data sharing and privacy and, upon consent, makes any blood leftover from clinical care eligible for BioVU banking. The VUMC Institutional Review Board oversees BioVU and approved this project. All data included in this study was de-identified and unlinked to any identifying information. This study was reviewed by the Vanderbilt University Medical Center IRB (IRB# 190418 and 170924) and designated as non-human subjects research. The research was conducted in accordance with the principles of the Declaration of Helsinki.

**CLHNS**: We thank the entire staff of the Office of Population Studies (OPS) Foundation in Cebu for their long-term work on the CLHNS. The CLHNS was supported by US National Institutes of Health grants R01DK078150, TW005596, HL085144; pilot funds from RR020649, ES010126, and DK056350; and the Office of Population Studies Foundation in Cebu. Additional support for data analysis was provided by US NIH R01DK072193.

**CHNS:** Support for CHNS is provided by NIH, R01HD30880 and R01AG065357. With additional funding from R01HD38700, R01 DK104371 and P30 DK056350, R01 HL108427, D43TW009077, 2CHD050924 and P30 AG066615, Additional support from China-Japan Friendship Hospital, Ministry of Health for support for CHNS 2009, Chinese National Human Genome Center at Shanghai since 2009, and Beijing Municipal Center for Disease Prevention and Control since 2011. We thank the National Institute for Nutrition and Health, China Center for Disease Control and Prevention, Beijing Municipal Center for Disease Control and Prevention, and the Chinese National Human Genome Center at Shanghai.

**Greenlandic Study**: We acknowledge all the participants and staff of the Greenlandic health surveys. The studies were approved by the Scientific Ethics Committee in Greenland (project 505-42, 505-95, project 2011–13 (ref. no. 2011–056978), project 2017-5582, project 2015–22 (ref. no. 2015-16426), and project 2021-09) and was conducted in accordance with the Declaration of Helsinki, second revision. All participants gave written informed consent.

**MESA**: The MESA projects are conducted and supported by the National Heart, Lung, and Blood Institute (NHLBI) in collaboration with MESA investigators. Support for the Multi-Ethnic Study of Atherosclerosis (MESA) projects are conducted and supported by the National Heart, Lung, and Blood Institute (NHLBI) in collaboration with MESA investigators. Support for MESA is provided by contracts 75N92020D00001, HHSN268201500003I, N01-HC-95159, 75N92020D00005, N01-HC-95160, 75N92020D00002, N01-HC-95161, 75N92020D00003, N01-HC-95162, 75N92020D00006, N01-HC-95163, 75N92020D00004, N01-HC-95164, 75N92020D00007, N01-HC-95165, N01-HC-95166, N01-HC-95167, N01-HC-95168, N01-HC-95169, UL1-TR-000040, UL1-TR-001079, UL1-TR-001420, UL1TR001881, DK063491, and R01HL105756. The authors thank the other investigators, the staff, and the participants of the MESA study for their valuable contributions. A full list of participating MESA investigators and institutes can be found at http://www.mesa-nhlbi.org. This study was also supported in part by the NHLBI contracts R01HL151855, R01DK081572 and U01HG011723.

**MGI**: The authors acknowledge the Michigan Genomics Initiative participants, Precision Health

at the University of Michigan, the University of Michigan Medical School Central

Biorepository, and the University of Michigan Advanced Genomics Core for providing data

and specimen storage, management, processing, and distribution services, and the Center

for Statistical Genetics in the Department of Biostatistics at the School of Public Health for

genotype data curation, imputation, and management in support of the research reported

in this publication.

**VA Million Veteran Program (MVP)**: This research is based in part on data from the Million Veteran Program, Office of Research and Development, Veterans Health Administration, and was supported by award 5I01BX003362 (PIs: Chang, Tsao). This publication does not represent the views of the Department of Veterans Affairs or the United States Government.

**Core Acknowledgements (May 2024)**

**MVP Program Office**

- Sumitra Muralidhar, Ph.D., Program Director

US Department of Veterans Affairs, 810 Vermont Avenue NW, Washington, DC 20420

- Jennifer Moser, Ph.D., Associate Director, Scientific Programs

US Department of Veterans Affairs, 810 Vermont Avenue NW, Washington, DC 20420

- Jennifer E. Deen, B.S., Associate Director, Cohort & Public Relations

US Department of Veterans Affairs, 810 Vermont Avenue NW, Washington, DC 20420

**MVP Executive Committee**

-	Co-Chair: Philip S. Tsao, Ph.D.

VA Palo Alto Health Care System, 3801 Miranda Avenue, Palo Alto, CA 94304

-	Co-Chair: Sumitra Muralidhar, Ph.D.

US Department of Veterans Affairs, 810 Vermont Avenue NW, Washington, DC 20420

-	J. Michael Gaziano, M.D., M.P.H.

VA Boston Healthcare System, 150 S. Huntington Avenue, Boston, MA 02130

-	Elizabeth Hauser, Ph.D.

Durham VA Medical Center, 508 Fulton Street, Durham, NC 27705

-	Amy Kilbourne, Ph.D., M.P.H.

VA HSR&D, 2215 Fuller Road, Ann Arbor, MI 48105

-	Michael Matheny, M.D., M.S., M.P.H.

VA Tennessee Valley Healthcare System, 1310 24th Ave. South, Nashville, TN 37212

-	Dave Oslin, M.D.

Philadelphia VA Medical Center, 3900 Woodland Avenue, Philadelphia, PA 19104

-	Deepak Voora, MD

Durham VA Medical Center, 508 Fulton Street, Durham, NC 27705

**MVP Co-Principal Investigators**

- J. Michael Gaziano, M.D., M.P.H.

VA Boston Healthcare System, 150 S. Huntington Avenue, Boston, MA 02130

- Philip S. Tsao, Ph.D.

VA Palo Alto Health Care System, 3801 Miranda Avenue, Palo Alto, CA 94304

**MVP Core Operations**

- Jessica V. Brewer, M.P.H., Director, MVP Cohort Operations

VA Boston Healthcare System, 150 S. Huntington Avenue, Boston, MA 02130

- Mary T. Brophy M.D., M.P.H., Director, VA Central Biorepository

VA Boston Healthcare System, 150 S. Huntington Avenue, Boston, MA 02130

- Kelly Cho, M.P.H, Ph.D., Director, MVP Phenomics

VA Boston Healthcare System, 150 S. Huntington Avenue, Boston, MA 02130

- Lori Churby, B.S., Director, MVP Regulatory Affairs

VA Palo Alto Health Care System, 3801 Miranda Avenue, Palo Alto, CA 94304

- Scott L. DuVall, Ph.D., Director, VA Informatics and Computing Infrastructure (VINCI)

VA Salt Lake City Health Care System, 500 Foothill Drive, Salt Lake City, UT 84148

- Saiju Pyarajan Ph.D., Director, Data and Computational Sciences

VA Boston Healthcare System, 150 S. Huntington Avenue, Boston, MA 02130

- Robert Ringer, Pharm.D., Director, VA Albuquerque Central Biorepository

New Mexico VA Health Care System, 1501 San Pedro Drive SE, Albuquerque, NM 87108

- Luis E. Selva, Ph.D., Director, MVP Biorepository Coordination

VA Boston Healthcare System, 150 S. Huntington Avenue, Boston, MA 02130

- Shahpoor (Alex) Shayan, M.S., Director, MVP PRE Informatics

VA Boston Healthcare System, 150 S. Huntington Avenue, Boston, MA 02130

- Brady Stephens, M.S., Principal Investigator, MVP Information Center

Canandaigua VA Medical Center, 400 Fort Hill Avenue, Canandaigua, NY 14424

- Stacey B. Whitbourne, Ph.D., Director, MVP Cohort Development and Management

VA Boston Healthcare System, 150 S. Huntington Avenue, Boston, MA 02130

**MVP Publications and Presentations Committee**

- Co-Chair: Themistocles L. Assimes, M.D., Ph. D

VA Palo Alto Health Care System, 3801 Miranda Avenue, Palo Alto, CA 94304

- Co-Chair: Adriana Hung, M.D.; M.P.H

VA Tennessee Valley Healthcare System, 1310 24^th^ Ave. South, Nashville, TN 37212

- Co-Chair: Henry Kranzler, M.D.

Philadelphia VA Medical Center, 3900 Woodland Avenue, Philadelphia, PA 19104

**Qatar**: The authors would like to acknowledge the Qatar BioBank (QBB) and the Qatar Genome Project (QGP).

**REGARDS**: This research project is supported by cooperative agreement U01 NS041588 co-funded by the National Institute of Neurological Disorders and Stroke (NINDS) and the National Institute on Aging (NIA), National Institutes of Health, Department of Health and Human Service. The content is solely the responsibility of the authors and does not necessarily represent the official views of the NINDS or the NIA. Representatives of the NINDS were involved in the review of the manuscript but were not directly involved in the collection, management, analysis or interpretation of the data. The authors thank the other investigators, the staff, and the participants of the REGARDS study for their valuable contributions. A full list of participating REGARDS investigators and institutions can be found at: <https://www.uab.edu/soph/regardsstudy/>.
